# Importation models for travel-related SARS-CoV-2 cases reported in Newfoundland and Labrador during the COVID-19 pandemic

**DOI:** 10.1101/2023.06.08.23291136

**Authors:** Zahra Mohammadi, Monica Cojocaru, Julien Arino, Amy Hurford

## Abstract

During the COVID-19 pandemic the World Health Organization updated guidelines for travel measure implementation to recommend consideration of a region’s specific epidemiological, health system, and socioeconomic context. As such, travel measure implementation decisions require region-specific data, analysis, and models to support risk assessment frameworks. From May 2020 to May 2021, the Canadian province of Newfoundland and Labrador (NL) implemented travel measures that required self-isolation and testing of individuals returning from out-of-province travel. We found that during the pandemic travel to NL decreased by 82%. Our best model was 135 times more likely to explain reported travel-related cases arriving in NL than a model where travel volume and infection data did not consider the Canadian jurisdiction of origin. To test an approach used in other studies, we formulated a model without considering the travel-related case data and found that this model performed very poorly. We conclude that importation models need to be supported with data describing the daily number of travel-related cases arriving in Canadian jurisdictions and daily travel volumes originating from each country and each Canadian province and territory. While there was some reporting of this information during the COVID-19 pandemic, these data were not consistently reported or easily accessible.

## 1 Introduction

On January 31, 2020, due to an outbreak of SARS-CoV-2 in Wuhan, China, the World Health Organization (WHO) advised other countries to expect SARS-CoV-2 cases and be prepared for outbreak containment, but measures that would restrict travel and trade were explicitly not recommended (Grépin et al, 2021; World Health Organization, 2020c). State Parties were to notify WHO within 48 hours of the public health rationale and justification of measures that would significantly interfere with international traffic (World Health Organization, 2020c). Despite broad consensus prior to the pandemic that during a public health emergency travel measures significantly impacting travel and trade should not be implemented, during the initial phase of the COVID-19 pandemic most countries implemented such travel measures (Grépin et al, 2021; Piccoli et al, 2022; Shoichet, 2020). The next two years saw substantial variation in the implementation and strictness of travel measures between (Piccoli et al, 2022) and within (Reddy et al, 2021; Studdert et al, 2020) countries. In July 2021, WHO provided updated recommendations stating that international travel-related measures should be ‘proportionate to the public health risk’ and adapted to a country’s ‘specific epidemiological, health system and socioeconomic context’, and recommended a risk-based approach (World Health Organization, 2021).

The inconsistent implementation of travel measures during the first two years of the pandemic may have been due to the low quality of evidence to support policy. Systematic reviews (Burns et al, 2021; Grépin et al, 2021) report that travel measures may have had a positive impact on infectious disease outcomes, and reduced and delayed imported SARS-CoV-2 cases from Wuhan, but that overall the quality of evidence was low. Most evidence was due to modelling studies with a lack of ‘real world’ data (Burns et al, 2021), with inconsistent parameter estimates and assumptions, and that overlooked the impact of undetected cases outside of China (Grépin et al, 2021).

To better support modelling studies with ‘real world data’, and to consider the specific context in which travel measures are applied, our analysis focuses on Newfoundland and Labrador (NL), a Canadian province that implemented travel measures during the COVID-19 pandemic. Our analyses contribute data, parameter estimates, and models to support decisions and inform best approaches to reporting during a pandemic. In particular, we note there are few data and guidelines to inform travel measure implementation within countries during a pandemic. We develop models that consider data describing travel volume arriving in NL, infection prevalence at a traveler’s origin, and reported travel-related cases in NL. Three studies have previously considered all of these data types (Arnold et al., 2024; McCrone et al, 2022; Yang et al, 2021), but many importation modelling studies have had to proceed without all of these data sources available; for example, Godin et al (2021) and Steyn et al (2021) do not consider travel volumes. We develop di”erent models to determine the impact of data gaps on the reliability of importation models. The data gaps we consider are: incomplete information on travel volumes, no data describing reported travel-related cases in the region of interest, no information on infection underreporting at travelers’ origins, and infection prevalence and travel volume data only available for large geographic areas.

Travel volume data is often incomplete. For example, the OpenSky database and the Official Aviation Guide are the travel volume data sources used in Russell et al (2021), and these sources report only travelers that arrive by air, and not by land or sea. Exclusion of travelers arriving by some travel modes is just one type of exclusion that occurs in travel volume data sources. Other exclusions can be types of workers or residents of the destination jurisdiction. We corrected the travel volume data for exclusions, and estimate how travel volumes to NL changed during the pandemic and given the enacted travel measures.

Several studies have estimated the epidemiological risk due to imported infections without any data reporting travel-related cases (Linka et al, 2020; Menkir et al, 2021; Milwid et al., 2024; Russell et al, 2021), and other studies have used data that does not distinguish between travel-related and community cases (Chinazzi et al, 2020; Costantino et al, 2020; Hincapie et al, 2022; Hossain et al, 2020; Wells et al, 2020). Data describing travel-related cases in travelers arriving in Canada were not consistently easily accessible to researchers during the pandemic. Post-arrival testing of travelers occurred in Canada, and these travel-related cases were sometimes reported on provincial and territorial public health websites, reported on by the media (Zhao et al., 2020), and compiled into accessible formats by volunteers (Berry et al, 2020, 2021). Travel-related case data could have been used to assess the accuracy of the predictions made in Milwid et al. (2024) describing the number of infections in international arrivals to Toronto Pearson, Montréal-Trudeau, Vancouver International and Calgary International airports. Travel-related case data could also have been used to assess the accuracy of the number of importations predicted to occur in Atlantic Canada and the territories made by Hincapie et al (2022). These travel-related case data exist, but were not considered by Hincapie et al (2022), so in Section A of the Supplementary Information we make this comparison.

To replicate the approach to importation modelling when travel-related case data is unavailable (i.e., Hincapie et al 2022; Milwid et al. 2024), we formulate a mechanistic model that considers the processes that give rise to travel-related cases, but is not fit to the travel-related case data. This mechanistic modelling approach estimates parameters from published studies as a method to overcoming the limitations arising due to travel-related case data being unavailable. The predictions of this mechanistic model are validated with the travel-related case data, which are completely independent of the mechanistic model’s parameterization. We choose this modelling approach to test whether the predictions of mechanistic models parameterized independently of travel-related case data (i.e., as occurs in Milwid et al. 2024 and Hincapie et al 2022) are accurate and to provide evidence for why reporting of travel-related cases is necessary during a pandemic. We also test whether importation models are less reliable if travel volume and infection prevalence data are aggregated as Canada or international, rather than considering each country, and Canadian province or territory of origin to inform the types of data that are needed to model importations and support decisions to implement travel measures during a public health emergency.

## 2 Methods

### 2.1 Background

Newfoundland and Labrador (NL; population: 510,550; Statistics Canada 2021) is the second smallest Canadian province and has few points of entry. Most non-resident travelers to NL visit the island of Newfoundland (93%, Government of Newfoundland Labrador 2018, population: 483,895 Statistics Canada 2021) and arrive by air to St. John’s International airport. From May 4, 2020 to June 30, 2021, the government of NL implemented travel measures that required non-residents to complete Travel Declaration Forms (TDFs) and self-isolate for 14 days after arrival. Rotational workers, NL residents working in other provinces, are a significant proportion of the NL workforce (Hewitt et al, 2018) and during the COVID-19 pandemic were subject to specific self-isolation requirements and testing regimes.

### 2.2 Data overview

#### 2.2.1 Travel volume to NL

We consider three data sources that report travel volumes: International Air Transit Authority (IATA) flight passenger data; TDFs completed by non-NL residents and other non-exempt individuals upon arrival to NL during the COVID-19 pandemic; and Frontier Counts (FC; Statistics Canada 2020-Statistics Canada 2021) completed at the Canadian border. The three sources of travel volume data have some overlap and di”erent limitations (Table 1), and when combined with tourism surveys from the Government of Newfoundland and Labrador (2020-2021), we were able to estimate correction factors to determine the ‘total travel volume’ arriving in NL from January 2019 to March 2020, and September 2020 to May 2021 (Supplementary Information, Section B.6), where ‘total travel volume’ includes arrivals by air, sea, and land ports of entry, and all traveler types including crew members, NL residents, and rotational workers. After estimating the total travel volume, we then stratified arriving travelers as regular travelers or rotational workers, and by travel origins. For travelers arriving from Canada, the origin can be any of the nine provinces (excluding NL) and the territories, where the three territories are considered as one because there is insufficient information on infection underreporting to separate each territory. For travelers arriving from international origins, we considered the eight countries comprising the most arrivals to NL and aggregated all remaining travelers into an ‘other countries’ category (see Section B.5 of the Supplementary Information for details). The provincial and federal travel measures that applied to travelers arriving in NL during the pandemic are summarized in Table 2.

**Table 1.** Limitations of travel volume data sources. International Air Transport Authority (IATA, *s* = 1), Travel Declaration Forms (TDF, *s* = 2), and Frontier Counts (FC, *s* = 3) report an origin (either Canada or international), but report travel volumes that exclude some travelers that might spread infections to NL residents. When exclusions or exemptions apply to particular travel modes (air, sea, or land) or traveler types (i.e., crew or NL residents) the value of the exclusions indicator variable, MODES or TYPE, is 1; and the indicator variable is 0 if this exemption does not apply. These indicator variables appear in equation B1, and the magnitude of the correction for the exclusion is given in Table B1 (see Section B in the Supplementary Information). The travel origin in TCAR reports (*s* = 4) was not reported, but the information in these reports was used to estimate the magnitude of the exclusions for the other data sources.

| s | Data Source | Time period | Pandemic | Travel modes excluded | $\mathbb{I}_{\text{MODES}} =$ | Origin | Traveler type exclusions | $\mathbb{I}_{\text{TYPE}} =$ |
| --- | --- | --- | --- | --- | --- | --- | --- | --- |
| 1 | International Air Transport Authority (IATA) | Jan 2019 - March 2020 (monthly) | before | land, sea | 1 | Canada and International | Canadian crew | 1 if Canada, 0 if International |
| 2 | Travel Declaration Forms (TDF) | Sep 2020 - May 2021 (daily) | during | none | 0 | Canada and International | NL residents, crew, and other exempt travelers | 1 |
| 3 | Frontier Counts (FC) | Jan 2019 - May 2021 (monthly) | before and during | none | 0 | International | None | 0 |
| 4 | Department of Tourism, Culture, Arts, and Recreation (TCAR) | Jan 2019 - Dec 2021 (monthly) | before and during | none | N/A | Unknown | NL residents | N/A |

**Table 2.** Federal (Canada) and provincial (Newfoundland and Labrador) travel measures from September 2020 to May 2021 (Canadian Institute for Health Information, 2022). ‘Line’ corresponds to the numbering in Fig. 1A.

| Line | Dates | Measure | Level |
| --- | --- | --- | --- |
| 1 | 2020-03-13 | Cruise ship season postponed | Fed |
|  | 2020-03-14 | 14-day self-isolation required for individuals returning from international travel | Prov |
|  | 2020-03-20 | 14-day self-isolation required for individuals returning from out-of-province travel | Prov |
| 2 | 2020-05-04 | Travel declaration forms and self-isolation plan required for non-NL resident entry to NL | Prov |
| 3 | 2020-06-09 | Relaxation of travel measures for foreign nationals with immediate family in Canada | Fed |
| 4 | 2020-07-03 | Atlantic bubble: No self-isolation requirement for residents of P.E.I., N.B. and N.S. | Prov |
| 5 | 2020-08-31 | Relaxation of travel measures for non-Atlantic Canada residents who own a second home or cabin in NL | Prov |
| 6 | 2020-10-20 | Relaxation of travel measures for international students attending institutions with a COVID-19 readiness plan | Fed |
| 7 | 2020-11-26 | Atlantic bubble suspended | Prov |
| 8 | 2021-02-01 | All international passenger flights must land either at the Vancouver, Calgary, Toronto or Montréal airports | Fed |
| 9 | 2021-03-27 | Passengers on provincial ferries limited to 50% of capacity | Prov |

#### 2.2.2 Infection prevalence at travelers’ origin

We estimate infection prevalence at the travelers’ origin from daily incidence in the Canadian provinces and territories as reported by the Public Health Agency of Canada (Canada) and John Hopkins University (international; Dong et al 2020) and adjusted for reporting delays as this information is used for explanatory variables in statistical models. We multiplied daily reported infections by an underreporting coefficient based on seroprevalence data reported by the COVID Immunity TaskForce (Canada) and using the approach described in Klamser et al. (2023) (international; see section C in the Supplementary Information for complete details). We estimate infection prevalence as a proportion by dividing by the population size.

#### 2.2.3 Travel-related cases reported in NL

Daily travel-related cases of Canadian and international origin were obtained from Newfoundland and Labrador Health Services - Digital Health (Figure B2 in the Supplementary Information; Newfoundland and Labrador Health Research Ethics Board reference number 2021.013), but were also reported publicly in Public Service Announcements from the NL Department of Health and Community Services during the pandemic. In NL, close contacts of travel-related cases were required to undergo asymptomatic testing, and if positive were reported as ‘close contacts of travelers’, and were not included in the reported number of travel-related cases. In this manuscript, travel-related cases arise from imported cases that test positive.

### 2.3 Modelling overview

#### 2.3.1 Statistical importation models

We formulated statistical models with a linear model structure. The statistical models were generalized linear models with a negative binomial or Poisson error distribution and where the response variable is either the daily travel-related cases reported in NL from Canada or from international origins. The statistical models have explanatory variables that are daily total travel volume from an origin (Section 2.2.1), daily infection prevalence at origin (Section 2.2.2), or both these variables and interaction terms. For models to predict daily travel-related cases of Canadian origin, we considered stratification of all model variables for each Canadian province and the territories, and aggregation of variables for all of Canada. For models to predict daily travel-related cases of international origin we consider stratification of all model variables for eight common countries of origin with all other countries aggregated as ‘other international’, and an aggregated model with all non-Canadian countries of origin combined. Models were fit using glm.nb from the MASS R package (Venables and Ripley, 2002) and prediction intervals were generated using add pi from the ciTools package (Haman and Avery, 2025).

#### 2.3.2 Model without travel-related case data

Using a modelling approach similar to that of Milwid et al. (2024) and Hincapie et al (2022), we consider the epidemiology of infection importation to predict the number of travel-related cases expected to be reported each day in NL from September 2020 to June 2021. The processes we consider include travel volumes, infection prevalence at a travelers’ origin, days since exposure for infected travelers, and testing policies in NL (Table D5 in the Supplementary Information contains descriptions of variables and parameters). Unlike the statistical models which are fit to the travel-related case data, this independently parameterized mechanistic model is developed based on the processes typically represented in mechanistic importation models, with the number of travel-related cases predicted based on the testing of travelers that occurred in NL during the pandemic. Figure D3 in the Supplementary Information is an overview of the mechanistic model.

For travelers arriving in NL, Polymerase Chain Reaction (PCR) tests were mandatory for anyone symptomatic, for travelers that were passengers on flights for which exposure notifications were issued, and for rotational workers upon their return to NL (described in Tables D6 and D7 of the Supplementary Information). For travelers from Canadian origins, the predicted number of travel-related cases reported in NL at *t* is,

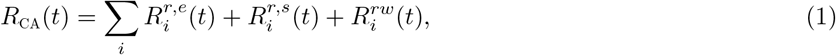

where the sum *i* is across all Canadian provinces (except NL) and the territories, 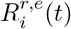 is the number of regular travelers that are predicted to test positive and are tested due to an exposure notification, 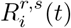 is the number of regular travelers that are predicted to test positive and requested a test because they developed symptoms postarrival and 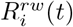 is the number of rotational workers that are predicted to test positive on at least one mandatory post-arrival test.

For travelers from international origins, the predicted number of reported travel-related cases in NL is,

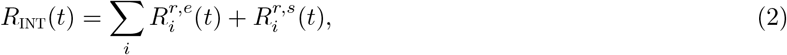

which is similarly defined as equation 1 except that the sum is across di”erent countries of origin, *i*, and only regular travelers are considered in the sum because by definition rotational workers do not return from international origins.

#### 2.3.3 Assessment of model fit

Model fit was assessed by calculating the negative log likelihood (nLL). The likelihood ratio test statistic is calculated as,

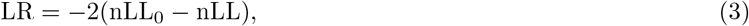

where nLL_0_ is a null model (or ‘constant only’ model) with the only fitted parameters the intercept, *ω*_0_, and the overdispersion parameter, *ε*, when the negative binomial error distribution is considered (see equations E22 and E25). The likelihood ratio test statistic was not calculated for the models that were not fit to travel-related case data (i.e., the mechanistic model) because these models were not nested.

The accuracy of the predictions for the model that did not consider the travel-related case data are assessed by calculating the likelihood of *R*_CA_(*t*) given the reported number of travel-related cases of Canadian origin (*Y*_*t*,CA_), and the likelihood of *R*_INT_(*t*) given the reported number of travel-related cases of international origin (*Y*_*t*,INT_), and assuming a Poisson error distribution, i.e.,

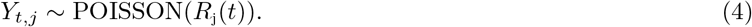

The Poisson distribution was used because it is not possible to fit an overdispersion parameter as the mechanistic model predicts only the mean number of reported travel-related cases and not the variance.

The best model is defined as the model with the smallest negative log likelihood. As the aim of our study is to most accurately model importations the reliability of the model is the only relevant consideration, however, our best models did have the most parameters and for context we also report the corrected Akaike Information Criteria (AICc) scores. Goodness of fit for predictions made by the best model was assessed by calculating the residual deviance and testing the null hypothesis that the predictions of the fully saturated model and the best model were di”erent.

The code and data used for analysis is available from Hurford, A. and Z. Mohammadi (2025).

## 3 Results

We found that the total travel volume arriving in NL declined by 82% during the pandemic while travel measures were enacted (September 2020 to May 2021) compared to the same period a year prior (Figure 1A; dark purple line). Relative to the same nine months one year prior to the pandemic, the average percentage of travelers arriving in NL increased from British Columbia (2.5 to 5.2%), Alberta (6.7 to 16%), the Canadian territories (0.3 to 3.6%), and international origins not the United States (8.2 to 15%; Figure 1E,F). The average percentage of travelers arriving in NL during the pandemic decreased from Ontario (27 to 21%), Québec (15 to 4.7%), and the United States (14 to 9.2%; Figure 1E,F).

**Fig. 1.**
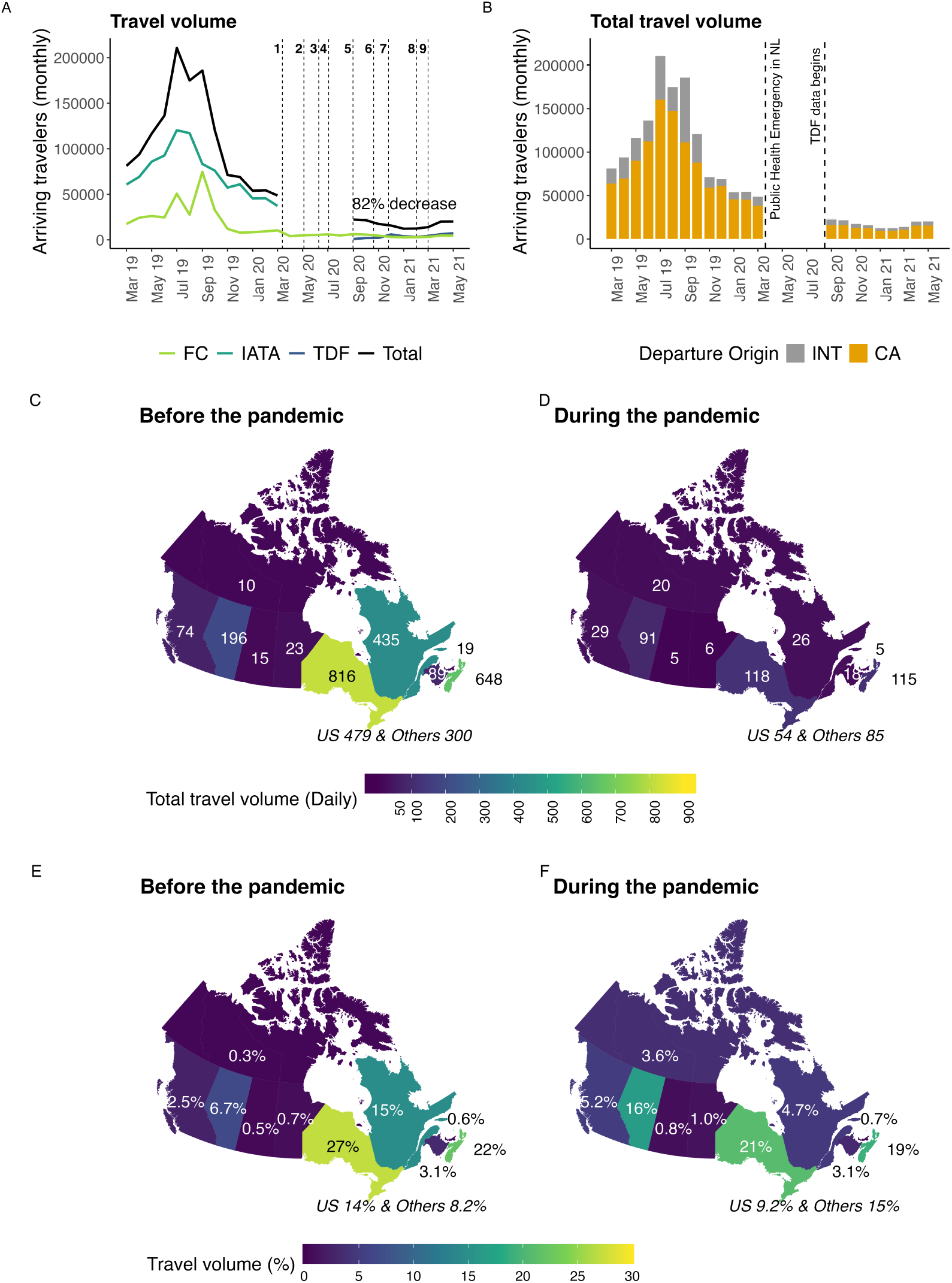
A) The total travel volume arriving in Newfoundland and Labrador before and during the pandemic (dark purple line, equation B1 in the Supplementary Information), and travel volumes reported by individual data sources: International Air Transport Authority (IATA, dark green line); Travel Declaration Forms (TDF; blue line); and Frontier Counts (FC; light green line). The numbered vertical lines correspond to travel measures implemented in NL (see Table 2). B) Total travel volume arriving from Canadian and international origins. C) and D) The average number of travelers arriving daily before and during the pandemic and their region of origin, where these averages are taken over nine months. E) The average percentage of arrivals from particular origins for nine months during the pandemic (September 2019 to March 2020, and April to May 2019), and F) for the same nine months during the pandemic (September 2020 to May 2021).

Our best statistical models had all explanatory variables and interaction terms, where all variables are stratified for each of the ten possible Canadian jurisdictions of origin, and nine international regions of origin. The best model predicting travel-related cases arriving in NL from Canada (Figure 2A) was 177.7 times more likely to explain the data than a null model consisting of only an intercept (Table 3). The residual deviance for the best model was 233 with 237 degrees of freedom. The chi-squared test found that the null hypothesis that the data are di”erent than the model predictions could not be rejected (*p* = 0.56). All the best models reported in Table 3 assume a negative binomial error distribution except the ‘Travel volume × infection prevalence (provinces)’ model, which could not be fit with a negative binomial error distribution and instead assumes the Poisson error distribution (see Section E.1 for further details).

**Table 3.** Model fit for predicted daily travel-related cases reported in NL from Canadian and international origins. Models where variables are stratified for Canadian provinces and the territories are denoted with ‘provinces’ in parenthesis. Models were variables are aggregated for the Canadian provinces and the territories or for all international origins are denoted with ‘aggregated’ in parenthesis. The number of parameters that are fit is *K*.

| Canada | nLL | LR | K | AICc |
| --- | --- | --- | --- | --- |
| Travel volume × infection prevalence (provinces) | 244.2 | 177.7 | 31 | 559.1 |
| Infection prevalence (provinces) | 270.8 | 124.8 | 12 | 566.8 |
| Travel volume (provinces) | 294.8 | 76.8 | 12 | 614.9 |
| Travel volume × infection prevalence (aggregated) | 311.7 | 43.1 | 5 | 633.6 |
| Travel volume (aggregated) | 321.4 | 23.6 | 3 | 648.9 |
| Infection prevalence (aggregated) | 326.6 | 13.1 | 3 | 659.4 |
| Constant only | 333.2 | 0 | 2 | 670.5 |
| Without travel-related cases | 538.2 | not nested | 0 | 1076.5 |
| International |  |  |  |  |
| Travel volume × infection prevalence (countries) | 131.4 | 42.5 | 28 | 325.7 |
| Infection prevalence (countries) | 143.0 | 19.4 | 10 | 306.8 |
| Travel volume × infection prevalence (aggregated) | 145.2 | 15.0 | 5 | 300.6 |
| Infection prevalence (aggregated) | 145.3 | 14.7 | 3 | 296.7 |
| Travel volume (countries) | 147.0 | 11.2 | 11 | 317.1 |
| Travel volume (aggregated) | 151.8 | 1.8 | 3 | 309.7 |
| Constant only | 152.7 | 0 | 2 | 309.4 |
| Without travel-related cases | 191.2 | not nested | 0 | 382.4 |

**Fig. 2.**
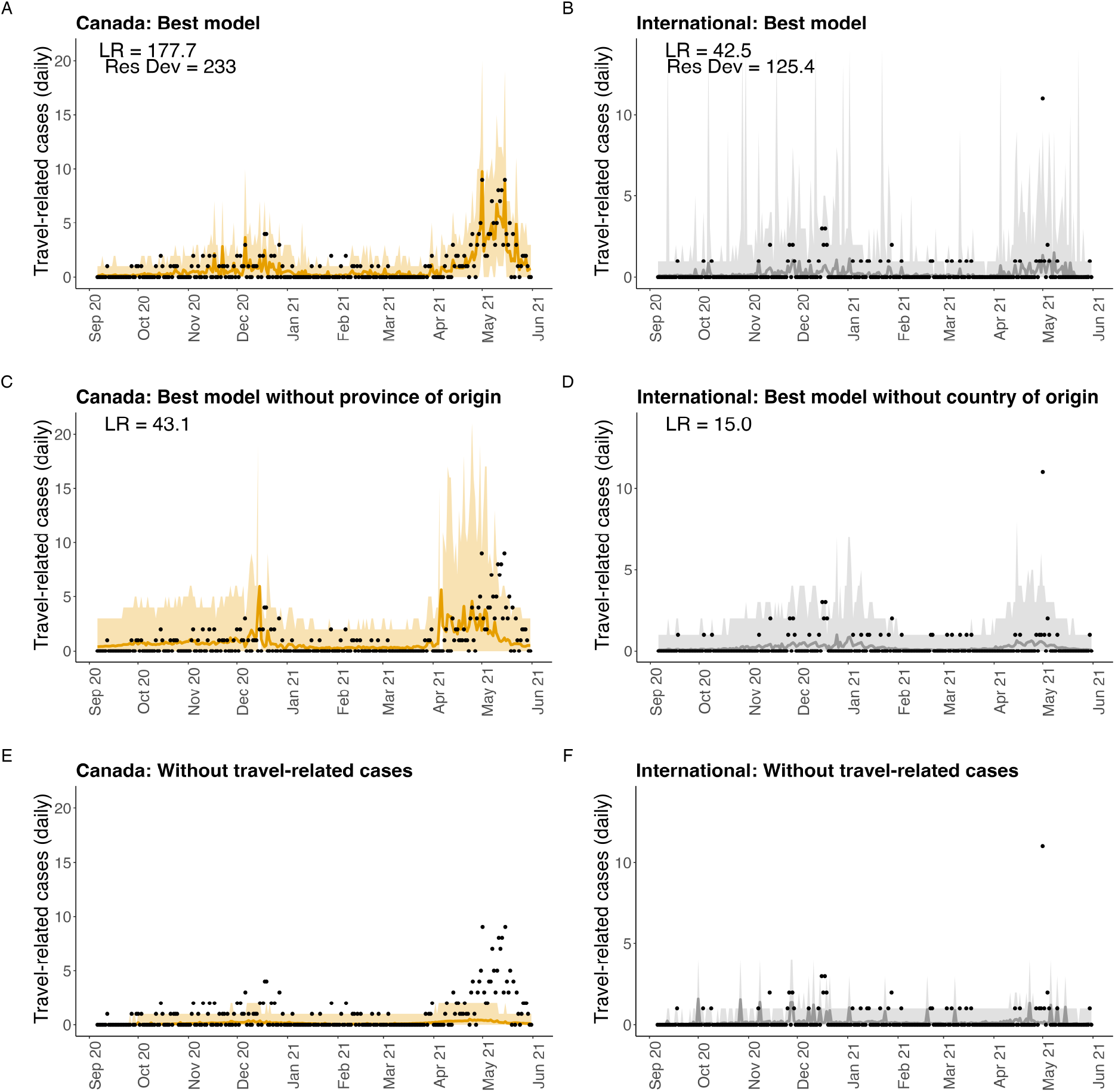
Reported travel-related cases as predicted by the best model, the best model with explanatory variables aggregated as Canada or international, and the model that was formulated independently of the travel-related case data (see Table 3 for the models). The shaded region is a 95% prediction interval and the travel-related case data are shown as black dots.

The best model that aggregated the explanatory variables without distinguishing the Canadian jurisdiction of origin was 43.1 times more likely to explain the data than the ‘constant only’ model consisting of only an intercept (Figure 2C). As such, the model that was stratified to consider each of ten Canadian jurisdictions of origin was 134.6 times more likely to explain the data than the best aggregated model. The mechanistic model that was not fit to the travel-related case data yielded worse predictions (nLL = 538.2; Figure 2E) than the model consisting of only an intercept and assuming a Poisson error distribution (nLL = 407.0; Table E8).

Travel-related cases arriving in NL from international origins were best predicted by a statistical model with all explanatory variables and interaction terms, and where all variables were stratified by each of nine possible origins (Figure 2B). The best model was 42.5 times more likely to explain the data than the ‘constant only’ null model (Table 3). The residual deviance of the best model was 125.4 with 241 degrees of freedom. The chi-squared test found that the null hypothesis that the data are di”erent than the model predictions could not be rejected (*p* = 1). When country-level data was not available the best model was 15.0 times more likely to explain the data than a constant model. The model that was not fit to the travel-related case data yielded worse predictions (nLL = 191.2; Figure 2F) than the model consisting of only an intercept and assuming a Poisson error distribution (nLL = 178.7; Table E8). The fitted coefficients for the best models are reported in Section E.1 of the Supplementary Information.

Only the TDF travel volume data source reports daily travel volume, so to determine the e”ect of using just one travel volume data source it was necessary to fit to the travel-related case data aggregated by month (nine observations). Results are reported in Table E9 of the Supplementary Information, however, generally, the lack of daily travel volume estimates (only one per month) reduced our ability to formulate and test the models. We also found that correcting infection prevalence for underreporting had only a small e”ect on model fit (Table E10 in the Supplementary Information).

## 4 Discussion

In July 2021, the World Health Organization (WHO) updated guidelines for international travel (World Health Organization, 2021) to advise that local epidemiology, and public health and health system performance and capacity should be considered to determine if travel measures are appropriate during a pandemic. Analysis of regional data and regionally-specific models are needed because decisions of whether travel measures are appropriate depend on regional characteristics.

We estimated that travel measures (described in Section B.2 of the Supplementary Information) reduced the total travel volume arriving in NL by 82% (September 2020 - May 2021) compared to the same nine months prior to the pandemic (Figure 1A, dark purple line). This finding is similar to another study finding an estimated 79% reduction in non-resident visitor volume to NL for 2020 compared to 2019 (Government of Newfoundland and Labrador, 2020). In other countries, air travel is estimated to have declined, on average, 63% for May 2020 (during the pandemic) as compared to May 2019 (before the pandemic; Russell et al 2021). The more substantial reduction in travel to NL may have been due to stricter travel measures for entry to NL than for other countries and regions during the study period. Most notably, non-NL residents entering NL from other regions of Canada were required to complete

Travel Declaration Forms (TDFs) and have a self-isolation plan to submit to a government representative at entry. These results help quantify the impact of travel measures implemented during a pandemic, which is necessary since modelling that predicts clinical cases depend on assumptions describing how travel measures a”ect travel volume (Hurford et al, 2021).

We found that during the pandemic there was a 10% increase in arrivals from Alberta and a 10% decrease in arrivals from Québec to NL (Figure 1E,F). Evidence for this result can be seen in Table B3 of the Supplementary Information, where the percentages before the pandemic (January 2019 - March 2020) are calculated from the International Air Transit Authority (IATA) data, and the percentages during the pandemic (September 2020 - May 2021) are calculated from the TDF data. This shift in the percentage of travelers arriving from di”erent provinces may be explained by the high percentage of NL’s rotational workers that work in Alberta (57%) as compared to Québec (2%; Table B3 in the Supplementary Information), while the travel volume of regular travelers from Québec before the pandemic is relatively high (11-26%, third highest behind Ontario and Nova Scotia, Table B3 in the Supplementary Information). Rotational workers likely continued working during the pandemic, while regular travelers may have delayed or canceled trips. Importation models used during public health emergencies often consider travel volume data. An implication of our findings is that the travel volume data used for these calculations needs to be reported daily, otherwise the type of importation modelling that can be done is very limited (i.e., cannot be stratified by province and territory of origin), and that travel volume data collected before the pandemic may not accurately represent the proportion of travelers arriving from di”erent travel origins. It is necessary to consider travel volume and infection prevalence estimates that are stratified by Canadian province and territory of origin as when these data are aggregated as ‘Canada’ the ability of the model to explain the data is substantially reduced.

Our results find that mechanistic importation models formulated independently of travel-related case data (i.e., Milwid et al. 2024 and Hincapie et al 2022) are much less reliable than the modelling that can be done, either as mechanistic or statistical models, when travel-related case data is available. The mechanistic modelling approach is to mathematically describe the processes that give rise to imported cases, and to estimate parameters from the published literature. The predictions of some mechanistic models (Hincapie et al, 2022; Linka et al, 2020; Nakamura and Managi, 2020; Russell et al, 2021) have not been tested or validated, and so the accuracy of this modelling approach is not known. It is perhaps unsurprising that the mechanistic models formulated without travel-related case data performed very poorly, yet travel-related case data is often unavailable to researchers that could use these data in their analyses. The aim of our study was to demonstrate that travel-related cases should be reported during a public health emergency because importation modelling is unreliable without these data. In that the constant only model performs better than these mechanistic models, travel-related case data is the most important data needed to support importation modelling because even a model with no explanatory variables is better than modelling that occurs without access to travel-related case data (Table 3). Without travel-related case data, the only type of modelling that is possible is mechanistic modelling, but mechanistic models can be fit to data, and if this had been done, mostly likely the mechanistic model would have performed similarly to the statistical models. As such, the mechanistic model performs poorly not because it is mechanistic, but because it is not fit to the travel-related case data.

Figure F4 in the Supplementary Information shows that it is plausible that the mechanistic model could fit the data better. The mechanistic model substantially underestimates travel-related cases (Figure 2E,F) and this could be because our data sources underestimate travel volumes and/or infection prevalence at travelers’ origin. This suggests that the mechanistic modelling could be improved with better travel volume data or recommending investment in better surveillance to determine infection prevalence. We found that information on underreporting did not improve our statistical models (see Section E.2.2 in the Supplementary Information), but accurate information on underreporting could be critical for mechanistic models (see Section F in the Supplementary Information). This conclusion further underscores the importance of travel-related case data as we would not know the mechanistic model was a substantial underestimate without being able to compare its predictions to the travel-related case data.

The processes that are described by mechanistic importation models may sound compelling, but there are a number of reasons why the assumptions of such models may not hold. Many studies consider only air travel volumes (Chinazzi et al, 2020; Hincapie et al, 2022; Nakamura and Managi, 2020; Russell et al, 2021; Wells et al, 2020), although travelers are likely to arrive via other travel modes and this proportion may change seasonally. No one travel volume data source reports the epidemiologically-relevant travel volume due to their scope and exemptions. Travel documents completed at international borders omit travelers that originate from within the country, and not all travel between countries require such documents to be completed (i.e., travel between countries in the European Union). Provinces may conduct surveys to understand tourist preferences, but such surveys (i.e., Government of Newfoundland and Labrador 2020-Government of Newfoundland and Labrador 2021) usually focus only on non-resident travel, while returning residents and workers are a potential source of imported infection. We combined four data sources (see Section B in the Supplementary Information) to overcome limitations of each travel volume data source and to estimate total travel volume; however, this was time-consuming, and to model importations urgently during a public health emergency, it is necessary that daily total travel volume data is available in real time.

While our best statistical models fits the reported travel-related case data well (Figure 2), the focus of our study was to understand the impacts of data gaps rather than to find the best importation model. The best importation modelling approach has been studied by others. Arnold et al. (2024) performs statistical modelling to describe the number of cases reported in international travelers isolating in government-managed quarantine facilities in New Zealand during the COVID-19 pandemic. In Arnold et al. (2024), countries of origin that have low numbers of arrivals, low numbers of cases, or both, are referred to as ‘low information countries’. In our study, the di”erences between the best statistical model’s predictions and the reported travel-related cases in NL are likely due to low numbers of cases and low travel volumes, particularly for travel-related cases of international origin. Nonetheless, the model fits shown in Figure 2 demonstrate that our model predictions have good correspondence with reported travel-related cases.

Regarding forecasting of imported infections arriving in a jurisdiction during a pandemic, the appropriate modelling approach depends on data availability. If travel-related case data reported in the destination jurisdiction are available, then statistical modelling is a good option. However, even when travel-related case data are available, it is good practice to develop both a more mechanistic model and statistical models, to contribute to our knowledge of how to best model importations, to better understand the processes that predict imported infections, and so we can avail of the strengths of each approach. A noted strength of mechanistic models, which is less reasonable for statistical models, is the ability to consider counterfactual scenarios (Ogden et al., 2024). Furthermore, while our analysis has demonstrated that travel-related case data, and travel volume and infection prevalence data available at the country, province, and territory level is necessary for reliable importation modelling, we did not have data that linked travel-related cases to the traveler’s origin. The availability of these linked data would further support the development and assessment of importation models to support public health decision-making during a pandemic and this information was available in Canada during the pandemic (Zhao et al., 2020), although not as easily accessible to researchers as was information on reported cases, vaccination levels, and variant prevalence, which was available for download from the Public Health Agency of Canada’s website.

Testing of travelers at points of entry is necessary to support early warnings for changes in epidemiological patterns (World Health Organization, 2022). Early in the COVID-19 pandemic, WHO situation reports described transmission classifications as imported cases only, sporadic cases, clusters of cases, local transmission, and community transmission where this information was self-reported by the State Parties (World Health Organization, 2020a,b). We do not consider it necessary for all regions to report the number of cases with a travel history (see also Martignoni et al 2024), but reporting of data describing the number of travel-related cases in a jurisdiction that is not experiencing widespread community transmission is necessary to support the development and refinement of importation models, and the results of our study definitively demonstrate this point. Generally, data, models, and analysis are needed to inform decisions to implement travel measures in a region, and our results aim to support decision-making and inform future approaches to model development.

## Data Availability

Data requiring ethics approval can be requested from the Newfoundland and Labrador Health Services, Digital Health. All publicly available data are archived online.

https://github.com/ahurford/importation-revision

## Acknowledgments

We thank three anonymous reviewers for comments that greatly improved the manuscript. All authors were supported by funding from Mathematics for Public Health (560523-2020), the One Health Modelling Network for Emerging Infections (560520-2020), and by the Emerging Infectious Disease Modelling Consortium funded by the Natural Sciences and Engineering Council of Canada. AH and JA are supported by the Canadian Network for Modelling Infectious Diseases (560516-2020). We thank the Newfoundland and Labrador Centre for Health Information (NLCHI; now Newfoundland and Labrador Health Services - Digital Health) and K. Lester for support in accessing the data.

## Declarations

### Ethics approval

Newfoundland and Labrador Health Research Ethics Board reference number 2021.013

### Availability of data and materials

Newfoundland and Labrador Health Services - Digital Health is the data custodian for the Travel Declaration Form data and Newfoundland and Labrador COVID-19 cases. Otherwise, data used for analysis is available from Hurford, A. and Z. Mohammadi (2025).

### Code availability

The code used for analysis is available from Hurford, A. and Z. Mohammadi (2025).

### Authors’ contributions

All authors conceptualized the study. ZM cleaned the data, completed the analysis, and made all the figures, although some analyses and figures were remade by AH to incorporate reviewer comments. ZM and AH interpreted the results and wrote the manuscript. All authors edited the manuscript.

