## Supplementary Material for "Importation models for travel-related SARS-CoV-2 cases reported in Newfoundland and Labrador during the COVID-19 pandemic"

### Appendix A An importation model that does not estimate parameters from travel-related case data

Hincapie et al (2022) predicts the number of importations arriving to different Canadian jurisdictions from January 1, 2020 to August 31, 2021, although this information is difficult to access given how these quantities are reported in their work (see Figures 3 and 4 in Hincapie et al 2022 right panels, vertical axis). For six of the seven jurisdictions Hincapie et al (2022) considers in their Figure 4 the provincial and territorial governments reported the value of the predicted quantity (the daily number of imported infections). For the number of cases that were travel-related from July 1, 2020 to May 31, 2021 for Newfoundland and Labrador, Nova Scotia, Prince Edward Island, New Brunswick, Northwest territories and Yukon see Hurford et al (2023). In that Hincapie et al (2022) predicts the number of importations for these jurisdictions without considering the number of reported importations, the motivation for our manuscript was to test whether such an approach to importation modelling, without considering travel-related case data, might be accurate.

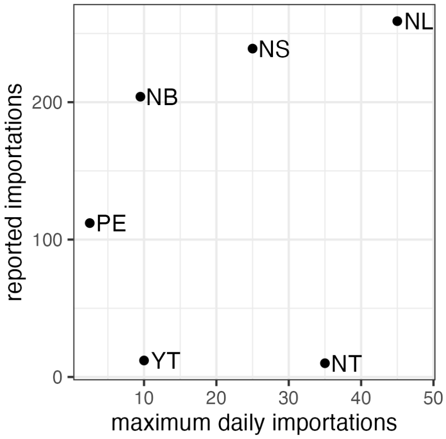


**Fig. A1** A comparison of the reported imported cases for Newfoundland and Labrador, Nova Scotia, Prince Edward Island, New Brunswick, Northwest territories and Yukon from July 1, 2020 to May 31, 2021 (Hurford et al, 2023) with the maximum number of daily importations predicted by the model of Hincapie et al (2022) for the period January 1, 2021 to August 31, 2021.

In assessing the predictions of the number of imported cases in Hincapie et al (2022), we first note that Northwest territories reported only 10 imported cases for the entire period July 1, 2020 to May 31, 2021 (Figure A1) while Figure 4 (right panels in Hincapie et al 2022) shows up to 35 imported cases per day. In the middle panel (Figure 4 of Hincapie et al 2022), the number of new cases (blue) for Northwest territories only reaches a maximum of around 5. Both these results suggest the importation modelling of Hincapie et al (2022) may very substantially overestimate the number of importations to Northwest territories.

The number of imported cases predicted by the Hincapie et al (2022) modelling for New Brunswick may be a substantial underestimate. For the Hincapie et al (2022) modelling the maximum number of daily importations for New Brunswick is around 9 importations per day, which is notable in that it is a lot less than this same quantity for the Nunavut (around 38 importations per day) and Northwest territories (around 35 importations per day; see Hincapie et al 2022, Figure 4 right panels vertical axis), but where these two Canadian territories are much smaller and more remote than the province of New Brunswick. Hurford et al (2023) reports New Brunswick as having the third most imported cases (204) from July 1, 2020 to May 31, 2021, with the Canadian territories of Yukon (12) and Northwest territories (10) reporting many fewer importations. A reason that the Hincapie et al (2022) modelling might underestimate importations to New Brunswick relative to other small Canadian jurisdictions is that a high proportion of travelers to New Brunswick may enter by vehicle through the land borders at Québec and the United States, whereas it may be more common to enter the other provinces of Atlantic Canada and the territories by air. The Hincapie et al (2022) importation modelling approach only considers arrivals by air and this may be the reason that their modelling substantially underestimaties imported infections to New Brunswick.

For an understanding of the epidemiological dynamics in Atlantic Canada and Canada's territories from July 1, 2020 to May 31, 2021, including reported imported cases, see Hurford et al (2021) as these results are based on data reported by the provincial and territorial governments rather than being model predictions as occurs in Hincapie et al (2022).

This detailed discussion of Hincapie et al (2022) has been presented to show the need for the analysis presented in our manuscript, which considers the impact of travel-related case data being unavailable to support importation modelling. Similar to Hincapie et al (2022), the work of Milwid et al. (2024) predicts the number of imported cases to Canadian airports, but the accuracy of these predictions is not known because data describing the number of infections reported in travelers arriving at these airports is not available. For this reason, the focus of our manuscript has been to assess whether it is possible to accurately forecast imported infections when travel-related case data is not available.

### Appendix B Travel volume to NL

In this section, we provide additional details describing the travel volume data that is used to determine the change in travel to Newfoundland and Labrador (NL) during the pandemic, and as is used in models (i.e., as explanatory variables in the statistical models).

#### B.1 Pre-pandemic

For the pre-pandemic period, we had available International Air Transport Association (IATA) travel data, which reports the number of passengers traveling in all classes for flights to and from NL from January 2019 through March 2020 (International Air Transport Association, 2020). We considered the origins and the destination (NL) that was reported for each trip (i.e., not layovers). The data was classified into three categories: inbound, outbound, and within province, and in our analyses we consider only the inbound air travel volume.

#### B.2 During the pandemic

##### B.2.1 Travel measures

We wish to understand the impact that COVID-19 measures had on travel to NL. In response to the COVID-19 pandemic in NL, several travel measures were implemented at the federal or provincial levels (summarized in Table 2). At the federal level, on March 16, 2020 entry into Canada by air was allowed only to Canadians and permanent residents of Canada, and citizens of the United States of America. Air crews, travelers arriving in Canada in transit to a third country, diplomats, or immediate family members of Canadian citizens were also exempt (Trudeau, 2020). At the provincial level, on March 20, 2020 14-day self-isolation was ordered for all individuals entering NL from outside the province (Exemption Order, 2020). This order included some exemptions, e.g. workers in transportation, essential workers, and rotational workers (Exemption Order 2, 2020). Further provincial travel measures were implemented on April 23, 2020 (Amendment No. 6, 2020). These measures required that all individuals arriving in NL from outside the province must complete a Travel Declaration Form (TDF) and have a self-isolation plan to submit to a government representative upon entry. Also, all individuals arriving in Labrador by motor vehicle from the province of Québec must immediately stop at their point of entry (indicated by a representative of the Government of NL) to submit their declaration form and their self-isolation plan (Amendment No. 8, 2020).

Effective May 4, 2020, all individuals were prohibited from entering NL except residents of Newfoundland and Labrador, asymptomatic workers, and individuals who received a travel exemption (Amendment No.11, 2020; Travel Exemption Order, 2020). There were limited numbers of exempted individuals who were not required to complete the TDF and submit the self-isolation plan. These were travelers who stayed in the province less than 24 hours, arrived daily or several times a day via the Labrador-Québec border, entered the province via the Labrador-Québec border for school reasons, or arrived weekly or several times a week in the province (Declaration Exemption Order, 2020; Self-Isolation Exemption Order, 2020). Submitting a TDF at the Labrador-Québec border relaxed further on June 25, 2020 especially for the residents of Labrador City, Wabush, Fermont, the Labrador Straits area, Blanc Sablon, and greater Québec Lower North Shore area (Labrador-Quebec Border Amendments, 2020). Completing a TDF was required even when the Atlantic Bubble was enacted (Atlantic Travel Amendments, 2020). All these travel orders were in place until the first phase of reopening. Effective on July 1, 2021, an approved reason to travel to the province, and completion of a TDF, was no longer required (Re-Opening - Travel, 2021). We use the TDF information compiled by the Newfoundland and Labrador Centre for Health Information (now Newfoundland and Labrador Health Services - Digital Health) to estimate arrival volume in the province during the pandemic.

##### B.2.2 Travel volume

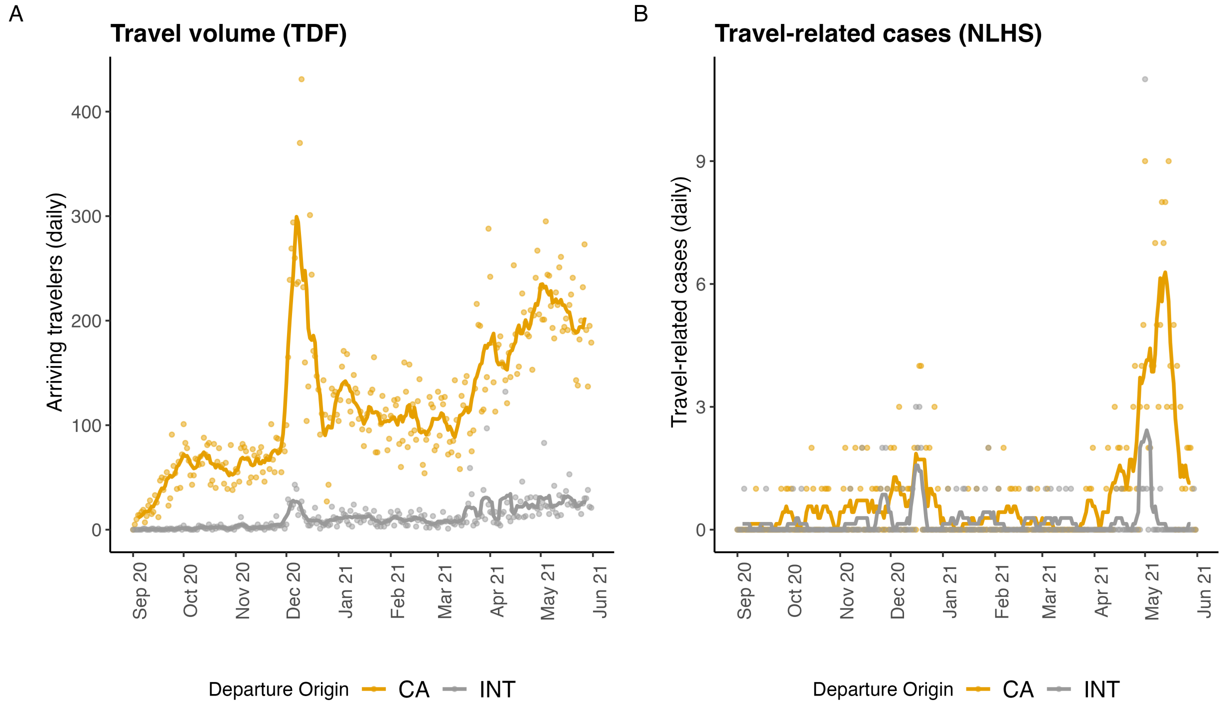


**Fig. B2** A. Travel volume of non-exempt travelers arriving in NL as reported by the TDF. Travel volume is a variable included in several of our models and may affect the number of travel-related cases reported in NL. B. Travel-related cases reported in the Newfoundland and Labrador Health Services (NLHS) - Digital Health COVID-19 database (September 2020 to May 2021; Canada - yellow; international - grey). Both daily values (dots) and 7-day rolling average (lines) are shown.

#### B.3 All time

Frontier Counts (FC) provide counts of entries into Canada by international travelers at Canadian ports of entry. Total travel volume includes all international travelers that were categorized as: Canadian residents returning to Canada, United States of America residents entering Canada, residents of countries other than the United States of America entering Canada and 'other' travelers which consist of foreign and resident crew members, diplomats, military personnel, immigrants and former residents (Statistics Canada, 2020-2021). For this study, we focus on NL ports of entry.

Data reported by Frontier Counts are extracted from different data sources provided by the Canada Border Services Agency (CBSA), i.e., Primary Inspection Kiosks (electronic systems at major airports), E311 Declaration Cards (forms completed at Canadian international airports without electronic system), and Telephone Reporting Centre-CANPASS (an electronic system for private plane or private boat or who report a land crossing by phone; Statistics Canada 2020-2021).

#### B.4 Estimating travel volumes not reported in the data sources

Travel volumes reported in different data sources have some travelers that are exempted (summarized Table 1) and our efforts to correct for these exemptions will be discussed in this section.

##### B.4.1 Non-air travel volume from Canadian origins

Some sources of travel volume data, such as IATA, report only arrivals by air. Reports from the NL Department of Tourism, Culture, Arts and Recreation (TCAR) from 2011 to 2018 indicate that air travel is the main mode of arrival for visitors to NL regardless of the season. The mean percentage of non-NL resident visitors arriving by air, auto, and cruise ships was 76%, 18%, and 6% respectively over three years 2016 to 2018 (Government of Newfoundland Labrador, 2017, 2018a).

From May to October 2016, 73% of non-NL residents arriving from Canadian origins to NL were by air (Government of Newfoundland Labrador, 2016); for arrivals from international origins this value was 85% (Government of Newfoundland Labrador, 2018), and with the remaining non-NL resident arrivals occurring by auto/ferry. Regardless of origin, the fraction of non-NL resident visitors who are using auto/ferry in spring (27%) was more than in fall (19%; Government of Newfoundland Labrador 2017).

Using all the TCAR reports from the Government of Newfoundland and Labrador (2020-2021), we estimated a correction factor, $\alpha_{CA,1}(t)$ (see Table B1 for notation), to calculate the total travelers (air and auto/ferry) originating from Canada. The estimated value multiplies the travel volume by air from Canadian origins and is: 1.24 (September, October, November, and March); 1.14 for the winter months (December, January, and February); and 1.37 for warmer months in spring and summer (April to August). For international arrivals, this correction for arrivals by auto/ferry is not necessary owing to the Frontier Counts data ($s = 3$), which reports arrivals by all travel modes.

**Table B1** Parameter values used to estimate the total travel volume arriving in Newfoundland and Labrador from Canadian ($i = CA$) and international ($i = INT$) origins. Data sources are: IATA ($s = 1$); TDF ($s = 2$); and FC ($s = 3$).

| Symbol | Description | Value |
|---|---|---|
| $Z_{i,s}(t)$ | Number of travellers arriving from origin $i$ as reported by data source $s$ | See Figure 1A. For the FC data ($s = 3$) we just have $i = INT$. |
| $\alpha_{i,s}(t)$ | Correction factor to estimate travellers by modes other than air | Values vary for different time periods with complete details in Table B3. $\alpha_{CA,1}(t)$ values are between 0.14 and 0.37, which means that travel volumes increase by 14-37% when the exclusions are considered. $\alpha_{INT,1}(t)$ was determined based on the FC data ($s = 3$) as this data reported arrivals for international origins for all modes. |
| $\beta_{i,s}(t)$ | For the IATA data ($s = 1$) excluded travellers are domestic crew members. For the TDF data ($s = 2$) exempt travelers are crew, NL residents, and other exempt travellers | Values vary monthly with complete details in Table B3. $\beta_{\mathrm{CA},1}(t)$ is between 2,107 and 24,952, $\beta_{\mathrm{CA},2}(t)$ is between 5,863 and 15,048, and $\beta_{\mathrm{INT},2}(t)$ is between 1,916 and 5,620 travellers excluded each month. |
| $h_i^{tot}(t)$ | Fraction of travellers from each Canadian province and territory | Values vary monthly with complete details in Table B3. |
| $h_i^{rw}$ | Fraction of rotational workers from each Canadian province or territory | Assumed not to vary over time with values reported in Table B3. |

##### B.4.2 Crew members and exempt travelers

Monthly crew member travel volume from international origins was obtained from the Frontier Counts data source ($s = 3$) both before and during the pandemic. The number of travelers per month that were crew members from Canadian origins is assumed to be 2.5 times the volume of crew members from international origins.

Travel declaration forms data ($s = 2$) reported arriving travel volume each day from Canadian or international origins during the pandemic. TDF data does not consider travelers who were exempt from completing the form (see Table 1 in the main text and Section B.2). We were able to estimate the correction for excluded travelers for each month and assumed an equal number of excluded travelers arrived each day during that month.

To estimate the number of exempt travelers from international origins, we compared the two data sources: TDF ($s = 2$) and IATA ($s = 3$). Since $Z_{\text{INT},3}$ included all international arrivals, the difference between this data source and $Z_{INT,2}$ is the number of international exempt travelers per month, $\beta_{\text{INT},2}(t)$.

Travelers from Canada exempt from filling out a TDF were interprovincial workers, NL residents, domestic crew members, and exempt travelers via Labrador-Québec border entry (see Section B.2). The correction applied to the TDF data source, where $\beta_{\text{CA},2}(t)$ accounts for the exclusion of both domestic crew members and other exempt travelers. To estimate the portion of other arriving exempt travelers (not crew), we assumed that the number of total exempt arrivals (residents and non-residents) was 2.4 times the number of exempt non-NL resident arrivals (see Aleman et al 2021 for a justification of this assumption), where the number of exempt non-NL resident arrivals from domestic origins is assumed to equal the number of exempt non-NL residents arriving from international origins per month. This value of 2.4 may be an overestimate, but this overestimate compensates for underestimation for other reasons such as challenges in submitting the TDF for travelers arriving by land.

#### B.5 Proportion and type of travelers arriving from a given origin

The TDF data was used to estimate the monthly percentage of travelers originating from each country (international travel volume) arriving to Newfoundland and Labrador during the pandemic (Table B2). Our analysis considers the eight countries that comprised at least 2.5% of international arrivals to NL in at least one of the three month periods: September to November 2020; December 2020 to February 2021; and March to May 2021. All other countries were aggregated into an 'other countries' category. The eight countries that were considered individually were: France, Guyana, India, Norway, Philippines, Spain, United Kingdom, and United States of America. Bangladesh, Suriname, and Thailand were not included due to a period with no travelers arriving which meant that model coefficients could not be estimated.

**Table B2** Percentage of quarterly international travel volume from origin countries. Only countries that account for at least 2.5% in one quarter are listed. St. Pierre and Miquelon are excluded.

| Country | Sept-Nov 2020 | Dec 2020 - Feb 2021 | March - May 2021 |
|---|---|---|---|
| Bangladesh | 0 | 3.0 | 2.7 |
| France | 1.8 | 1.7 | 3.9 |
| Guyana | 0.6 | 2.8 | 1.6 |
| India | 0.6 | 2.9 | 2.1 |
| Norway | 5.5 | 2.9 | 1.8 |
| Philippines | 0.6 | 2.6 | 2.8 |
| Spain | 1.8 | 3.3 | 2.4 |
| Suriname | 0 | 0.7 | 5.2 |
| Thailand | 0 | 0.1 | 3.6 |
| UK | 15.8 | 9.3 | 6.3 |
| USA | 47.3 | 31.9 | 33.6 |

The percentage of travel volume for each Canadian province or the territories from the IATA (before the pandemic) and TDF (during the pandemic) data was used to estimate the percentage of regular travelers arriving from each Canadian province and the territories (Table B3).

The percentage of NL residents that are interprovincial employees working in each Canadian province or the territories is reported by Hewitt et al 2018. In this report, the largest number of NL residents that are interprovincial employees work in Alberta (57%), followed by Ontario (15%) and Nova Scotia (8%). These values are used to estimate the within-Canada origin of rotational workers (Table B3).

**Table B3** Values for the parameters in Table B1. $h_i^{tot}(t)$ (per month) is the fraction of Canadian travel volume that originates from each individual province or the territories. $\alpha_{\text{CA},1}(t)$ is the correction factor to estimate the number of travelers arriving from Canada by modes other than air before the pandemic. $\beta_{\text{CA},1}(t)$ is the number of excluded travelers for data source $s = 1$ per month (pre-pandemic) whereas $\beta_{\text{CA},2}(t)$ and $\beta_{\text{INT},2}(t)$ are the monthly number of Canadian and international excluded travelers from data source $s = 2$ (during the pandemic). $h_i^{rw}$ is the fraction of rotational workers from each Canadian province and territory.

| $h_i^{tot}(\%)$ | | | | | | | | | | | $\alpha_{\text{CA},1}(t)$ | $\beta_{\text{CA},1}(t)$ | $\beta_{\text{CA},2}(t)$ | $\beta_{\text{INT},2}(t)$ |
|---|---|---|---|---|---|---|---|---|---|---|---|---|---|---|
| Date | ON | QC | MB | NB | AB | NS | BC | SK | PEI | TR | CA | CA | CA | INT |
| 2019-01 | 31.69 | 24.35 | 0.58 | 3.5 | 8.82 | 27.15 | 2.16 | 0.52 | 1.03 | 0.2 | 0.14 | 2920 | | |
| 2019-02 | 32.05 | 25.84 | 0.68 | 3.79 | 7.52 | 26.88 | 2 | 0.44 | 0.61 | 0.21 | 0.14 | 2417 | | |
| 2019-03 | 33.07 | 24.07 | 0.75 | 3.73 | 6.99 | 27.97 | 2.29 | 0.29 | 0.49 | 0.35 | 0.24 | 3642 | | |
| 2019-04 | 33.8 | 22.96 | 0.72 | 3.97 | 7.72 | 27.03 | 2.09 | 0.6 | 0.75 | 0.36 | 0.37 | 3150 | | |
| 2019-05 | 37.07 | 17.07 | 1.07 | 3.7 | 7.88 | 28.66 | 2.64 | 0.87 | 0.69 | 0.34 | 0.37 | 4750 | | |
| 2019-06 | 42.03 | 13.73 | 1.49 | 3.23 | 9.74 | 23.9 | 4.1 | 0.58 | 0.68 | 0.53 | 0.37 | 5702 | | |
| 2019-07 | 45.19 | 11.03 | 1.37 | 2.7 | 13.31 | 20.21 | 4.3 | 0.94 | 0.54 | 0.41 | 0.37 | 14075 | | |
| 2019-08 | 44.14 | 11.81 | 1.17 | 2.83 | 10.7 | 22.29 | 4.67 | 1.22 | 0.73 | 0.45 | 0.37 | 6650 | | |
| 2019-09 | 38.38 | 16.25 | 1.5 | 3.55 | 8.26 | 26.09 | 3.76 | 0.92 | 0.93 | 0.36 | 0.24 | 24952 | | |
| 2019-10 | 33.86 | 19.34 | 1.1 | 3.33 | 9.14 | 28.6 | 2.9 | 0.47 | 0.73 | 0.52 | 0.24 | 9395 | | |
| 2019-11 | 33.32 | 18.8 | 0.55 | 3.99 | 8.1 | 30.33 | 2.84 | 0.52 | 1.1 | 0.44 | 0.24 | 2842 | | |
| 2019-12 | 34.95 | 14.93 | 0.9 | 4.05 | 10.99 | 27.68 | 4.4 | 0.59 | 0.8 | 0.7 | 0.14 | 2445 | | |
| 2020-01 | 35.01 | 16.69 | 0.78 | 4.39 | 9.57 | 28.1 | 3.87 | 0.62 | 0.7 | 0.27 | 0.14 | 2107 | | |
| 2020-02 | 33.33 | 19.82 | 0.78 | 4.44 | 7.73 | 28.01 | 4.03 | 0.42 | 0.93 | 0.52 | 0.14 | 2412 | | |
| 2020-03 | 33.75 | 16.37 | 0.91 | 3.97 | 7.17 | 31.26 | 4.43 | 0.62 | 0.85 | 0.67 | 0.24 | 2252 | | |
| 2020-09 | 23.78 | 3.72 | 0.72 | 4.45 | 9.31 | 52.64 | 2.38 | 0.52 | 1.24 | 1.24 | | | 15048 | 5620 |
| 2020-10 | 28.09 | 3.4 | 0.66 | 4.57 | 14.47 | 41.14 | 3.25 | 1.02 | 1.52 | 1.88 | | | 13838 | 4669 |
| 2020-11 | 25.7 | 4.86 | 0.69 | 5.49 | 16.38 | 39.38 | 4.81 | 0.54 | 0.74 | 1.42 | | | 10827 | 3399 |
| 2020-12 | 28.62 | 5.57 | 2.26 | 3.99 | 24.29 | 17.04 | 9.54 | 1.08 | 1.41 | 6.2 | | | 6941 | 2231 |
| 2021-01 | 30.1 | 7.17 | 1.16 | 5.27 | 22.84 | 20.55 | 7.01 | 0.69 | 0.61 | 4.61 | | | 5863 | 1916 |
| 2021-02 | 26.92 | 7.22 | 1.93 | 3.16 | 27.36 | 16.16 | 8.25 | 1.03 | 0.79 | 7.18 | | | 6548 | 2144 |
| 2021-03 | 28.41 | 10.07 | 1.9 | 3.15 | 25.88 | 12.75 | 8.74 | 1.33 | 0.68 | 7.1 | | | 6831 | 2238 |
| 2021-04 | 28.02 | 7.2 | 1.47 | 3.49 | 26.15 | 15.59 | 9.21 | 1.16 | 0.71 | 7 | | | 9845 | 3096 |
| 2021-05 | 28.18 | 6.98 | 1.53 | 3.58 | 27.92 | 14.68 | 8.37 | 1.67 | 1.11 | 5.98 | | | 9065 | 2841 |
| $h_i^{rw}(\%)$ | 15 | 2 | 1 | 4 | 57 | 8 | 4 | 2 | 2 | 5 | | | | |

#### B.6 Estimating total travel volume

We estimated the total travel volume (where 'total travel volume' includes arrivals by air, sea, and land ports of entry, and all traveler types including crew members, NL residents, and rotational workers) arriving in NL by considering three data sources: International Air Travel Association data ($s = 1$), Travel Declaration Forms ($s = 2$), and Frontier Counts ($s = 3$). These data are each with limitations, and each reported for different time periods (Table 1). We estimate the total travel volume considering all travel modes, travelers and crew, from the origin $i$ at time $t$ by introducing indicator variables that correct for exclusions in the data sources,

$$V_{i,s}(t) = (1 + \alpha_{i,s}(t)\mathbb{1}_{\text{MODES}})Z_{i,s}(t) + \beta_{i,s}(t)\mathbb{1}_{\text{TYPE}}, \tag{B1}$$

where $Z_{i,s}(t)$ is the number of travelers arriving from origin $i$ per day ($s = 2$) or per month ($s = 1, 3$) reported from a data source, $s$. Possible origins are international ($i = \text{INT}$) or Canada ($i = \text{CA}$), but in subsequent calculations below we will further partition departures from Canada into each non-NL Canadian province and the territories (where all three Canadian territories are combined) and international into arrivals from one of the eleven countries or an 'other' category that aggregates all other countries.

The indicator variable $\mathbb{1}_{\text{MODES}}$ applies to data that does not report arrivals by all modes, and applies to the IATA data ($s = 1$) because this travel volume reports only arrivals by air. The correction factor, $\alpha_{j,s}(t)$, multiplies the reported travel volume by air. The indicator variable $\mathbb{1}_{\text{TYPE}}$ applies when some individuals are excluded from the reported travel volume, and the correction is additive. This correction applies to the IATA data, $\beta_{i,1}(t)$, for crew members that are not reported, and to the TDF data, $\beta_{i,2}(t)$, because arriving crew members on flights, ships, commercial freighters, NL residents, and other exempt travelers are not reported in this data.

The travel volume, $V_{i,s}(t)$, corrected for exclusions can be calculated for all data sources $s$; however, in practice the calculated values do not have a lot of overlap because the IATA data only corresponds to pre-pandemic time periods, the TDF data only corresponds to during the pandemic, and the FC data only reports travelers of international origin.

In equation B1, $i$ refers to the Canadian or international origin of travel, but both the IATA and TDF data report the Canadian province or territory of origin. Therefore, we estimate travel volume from different Canadian provinces and the territories as,

$$v_{i,s}(t) = V_{\mathrm{CA},s}(t) h_i^{tot}(t), \tag{B2}$$

where $h_i^{tot}(t)$ is the fraction of all Canadian travelers originating from each of the different non-NL provinces or the territories $i \in \{\mathrm{BC, AB, SK, MB, ON, QC, NB, NS, PE, TR}\}$ on a given date $t$, where Yukon, Northwest territories, and Nunavut are combined and denoted as TR, $s = 1$ corresponds to the IATA data when we are considering times before the pandemic, and $s = 2$ corresponds to the TDF data when we are considering times during the pandemic.

During the public health emergency in NL, different travel restrictions were applied to rotational workers. Rotational workers are NL residents who work in other provinces. The number of rotational workers entering NL is difficult to estimate, but Martignoni et al (2022) estimates that 6,000 NL residents are rotational workers. If we assume a rotational worker has a set schedule of two weeks of work and home (alternating), then approximately 200 rotational workers will enter NL each day. The fraction of rotational workers that work in any given province or the Canadian territories, $i \in \{\mathrm{BC, AB, SK, MB, ON, QC, NB, NS, PE, TR}\}$, is $h_i^{rw}$, was is estimated from Hewitt et al (2018) (Table B3). Therefore, the number of rotational workers arriving in NL during the pandemic from each of the different provinces and territories is,

$$v_i^{rw}(t) = 200 h_i^{rw}. \tag{B3}$$

The number of regular travelers (defined as all individuals that are not rotational workers) arriving from a Canadian origin, $i$, during the pandemic is,

$$v_i^{r}(t) = v_{\mathrm{CA},2}(t) - v_i^{rw}, \tag{B4}$$

where the travel volume is estimated from the TDF data ($s = 2$) because this data source corresponds to during the pandemic. The quantity $v_i^{rw}(t)$ does not change with time, but $v_i^{r}(t)$ does, so the dependence on time is written so that the compacted notation $v_i^{k}(t)$ can be used. We define

$$v_i^{r}(t) = v_{\mathrm{INT},2}(t), \tag{B5}$$

as the volume of regular travelers arriving from international origins during the pandemic, such that $v_i^{k}(t)$ is the volume of travelers arriving during the pandemic at time, $t$, that are rotational workers ($k = rw$, $i = \mathrm{CA}$), regular travelers arriving from a Canadian origin ($k = r$, $i = \mathrm{CA}$), or from an international origin ($k = r$, $i = \mathrm{INT}$).

### Appendix C Infection prevalence at origin

We formulated models to predict daily reported travel-related cases in NL that consider infection prevalence at a traveler's potential jurisdiction of origin. This section describes the data sources and calculations performed on these data.

#### C.1 Data sources

The data source for new SARS-CoV-2 cases (incidence) was the Public Health Agency of Canada for the Canadian provinces and territories (Public Health Infobase, 2020-2021). We used the daily infection incidence from the Center for Systems Science and Engineering (CSSE) at Johns Hopkins University (Dong et al, 2020) for incidence in the eight specific countries that comprised at least 2.5% of all international travelers during at least one three month period during our study (and no periods with 0%).

We used the method described in Martignoni et al (2023) to estimate the coefficient of underreporting for COVID-19 in region $i$, by considering the cumulative percentage of the population that was seropositive for SARS-CoV-2 antibodies relative to the number of reported COVID-19 cases in a region, $i$. We assumed the underreporting coefficient, $u_i$, was different in 2020 versus 2021 to be consistent with the methods we used for international cases. The cumulative percentage of the population infected between September 1, 2020 and December 31, 2020 is estimated from seroprevalence data (Blood Donation Organizations, 2023; Centers for Disease Control and Prevention, 2020-2021) and divided by the cumulative reported cases for each region thirteen days earlier because there are nine to twelve days between symptom onset and seroconversion (Lou et al, 2020). The underreporting coefficient, $u_i$, is calculated as the difference in the percentage of the population infected as estimated by the seroprevalence divided by the difference in the percentage of the population reported as infected (Table C4). Because more infections should have occurred than were reported we set the value of $u_i$ equal to 1 if the calculation results in a value less than 1.

We used the estimate of underreporting from Klamser et al. (2023) to correct reported infection prevalence for each of the eight countries comprising at least 2.5% of all international travelers during at least one three month period during our study (and now periods with 0%).

**Table C4** Underreporting coefficients for COVID-19 cases

| Country | July-Dec 2020 | Jan-June 2021 |
|---|---|---|
| British Columbia | 1.05 | 1.55 |
| Alberta | 2.02 | 2.56 |
| Saskatchewan | 1.38 | 2.80 |
| Manitoba | 3.09 | 1.00 |
| Ontario | 1.0 | 2.6 |
| Québec | 1.0 | 2.67 |
| New Brunswick | 1.0 | 1.0 |
| Nova Scotia | 1.0 | 1.0 |
| Prince Edward Island | 1.0 | 1.0 |
| Territories | 1.0 | 1.0 |
| Country | July-Dec 2020 | Jan-June 2021 |
| France | 2.1 | 1.4 |
| Guyana | 13.1 | 10.6 |
| India | 28.9 | 29.9 |
| Norway | 2.1 | 1.2 |
| Philippines | 28.5 | 26.2 |
| Spain | 2.2 | 1.6 |
| UK | 2.6 | 1.4 |
| USA | 2.6 | 1.9 |

### Appendix D Details of the model that does not consider travel-related cases

The mechanistic model aims to predict the number of reported travel-related cases arriving in NL in absence of this information being available to use in the modelling. We chose this approach to illustrate the value of travel-related case data being available to model importations. The mechanistic model is a linear model considering infection prevalence at a traveler's origin, travel volume from an origin to NL, the number of days since exposure for the infected traveler when arriving in NL, and the procedure for testing in NL after arrival. Figure D3 gives an overview of the model. The mechanistic model's parameters, which were estimated from the literature or assumed are provided in Table D5.

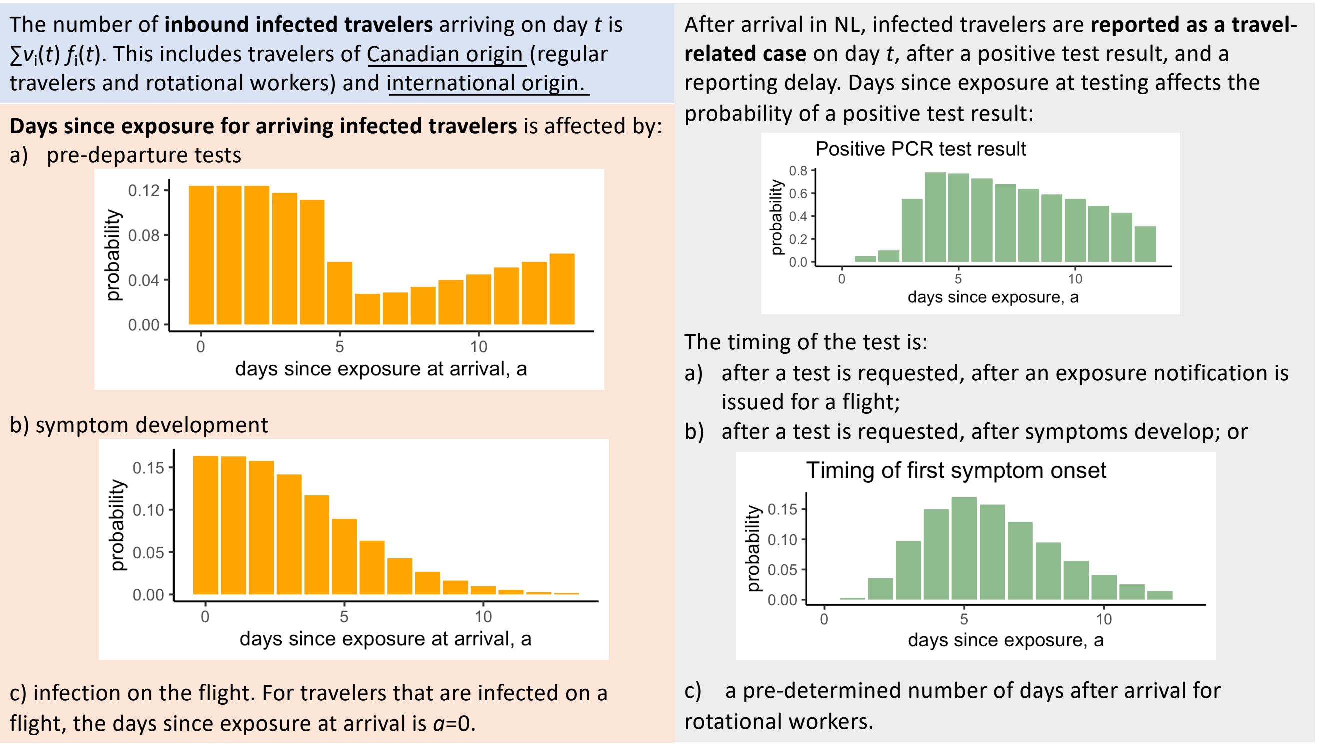


**Fig. D3** An overview of the mechanistic model, which is a modelling approach that can be used when travel-related case data is not available. The number of inbound infected travelers arriving on day $t$ is the product of $v_i(t)$: the number of travelers arriving from $i$ on day $t$, and $f_i(t)$: the prevalence proportion at $i$ on day $t$, summed over all travel origins, $i$. The days since exposure for infected travelers that arrive in NL depends on: a) positive pre-departure test results when such tests are required, b) the probability that symptom onset occurs before departure, and c) if the traveler was infected on the flight. Given a) and b) the distribution of days since exposure in arriving travelers are shown. Lastly, to be reported as a travel-related case in NL, the traveler must test positive on post-arrival test administered in NL. See Section D in the Supplementary Information for further details.

#### D.1 Estimating prevalence from incidence data

To estimate prevalence at a traveler's origin, we first estimate the incidence proportion (number of new cases divided by the population size) at a traveler's origin, $f_i(t)$, on a day $t$, as

$$f_i(t) = \frac{c_i(t + t_d)u_i}{N_i}, \tag{D6}$$

where $c_i(t)$ is the number of new cases reported on day $t$ at the origin $i$, $t_d = 11$ days is the delay between exposure and reporting (we note that Hendy et al 2021 reports this delay can be over two weeks), $N_i$, is the population size for each origin, $i \in \{\text{INT, BC, AB, SK, MB, ON, QC, NB, NS, PE, TR}\}$ (based on 2021 values estimated from Statistics Canada 2021 and The United States government 2021), and $u_i$ is an origin-specific correction factor to account for underreporting.

We convert from incidence proportion to prevalence proportion (the proportion of the population that have infections at time $t$) because we assume that the probability a traveler has an active infection at departure is equal to infection prevalence proportion at their origin. We assume that individuals have active infections for 14 days and approximate point prevalence as $14f_i(t)$. This calculation is not exact. Indeed, suppose prevalence is increasing until time $t$. Then at time $t$, more new infections than recoveries are occurring, so that point prevalence is actually more than $14f_i(t)$; the situation is reversed for decreasing incidence.

**Table D5** Variables and parameters that are estimated from the published literature or assumed for the mechanistic model

| Symbol | Definition | Value | Reference |
|---|---|---|---|
| $t$ | Days relative to a reference date | $\geq 0$ days | |
| $t_e$ | Days after arrival when an exposure notifications was issued | 3 days | Assumed. Ranged between 2 and 7 days |
| $t_r$ | Days between requesting a post-arrival test and the test occurring (for travellers without scheduled tests) | 3 days | Assumed |
| $t_{rep}$ | Days between testing and reporting in NL | 1 day | Assumed |
| $t_1$, $t_2$, $t_3$ | Days after arrival for the first, second, and third mandatory post-arrival test (if applicable) | | see Table D6 in the Supplementary Information for details |
| $a$ | Days since exposure of an infected person | 0 to 13 days | after 13 days probability of infection spread is negligible Ferretti et al. (2020) |
| $\bar{a}$ | Average number of days after arrival when symptoms first occur | 1 day | See Eq. (D13). The exact value is 1.49 days |
| $\lambda(a)$ | Probability of first developing symptoms given exposure $a$ days ago | Eq. (D10) | Lauer et al (2020) |
| $\gamma(a)$ | Probability of a true positive given exposure $a$ days ago | Eq. (D15) | Hellewell et al (2021) |
| $\psi$ | Fraction of travelers who travel despite having symptoms | 0.75 | Smith et al (2020) |
| $\rho$ | Fraction of infections that are symptomatic | 0.69 | Godin et al (2021) |
| $\sigma$ | Probability that symptomatic regular travellers get tested in NL | 0.8 | Assumed |
| $\phi_1$ | Probability a traveler is infected during an exposure subject to an exposure notification | 0.01 | Assumed |
| $\phi_2$ | Proportion of travelers arriving on day $t$ subject to the exposure notification | 0.05 | Assumed |

#### D.2 Infection status of arriving travelers

The infection status of arriving travelers indicates is the number of days since exposure. This quantity if used in the mechanistic model, for example, to estimate the probability of a positive PCR test.

We assume that the probability that a traveler is infected is equal to the prevalence proportion at the travelers origin, $f_i(t)$. We assume that the distribution of days since exposure of infected travelers at departure is uniformly distributed between 13 and 0 days ago, i.e.,

$$n_i(t,a) = \begin{cases} 14\frac{f_i(t)}{14} = f_i(t) & \text{for } a = 0,1,2,\ldots,13 \\ 0 & \text{otherwise,} \end{cases} \tag{D7}$$

where $a$ is the number of days since exposure or the 'age of infection'. Again, this is an approximation because if incidence is increasing at time $t$, there will be more individuals with recent exposures, so that the distribution of age of infection is right-skewed (i.e., the tail is to the right as there relatively fewer individuals exposed many days ago). An approximation is used because estimating the distribution of the ages of infection at any time from reported data is difficult. Indeed, whether a case is reported depends on the age of the infection: tests are requested once symptoms develop, test results can be false negatives, and both of these processes depend on the age of infection. Due to the development of symptoms and pre-arrival testing, not all infected travelers end up traveling. If a pre-arrival test was required, the number of infected travelers arriving from an origin $i$ that were exposed $a$ days ago is,

$$\mathrm{T}_i^{k,1}(t,a) = v_i^k(t)(1-\gamma(a-2))n_i(t,a), \tag{D8}$$

where $v_i^k(t)$ is the travel volume from origin $i$ of rotational workers ($k = rw$) or regular travelers ($k = r$) who departed on date $t$ (see equations B3, B4 and B5), $1-\gamma(a-2)$ is the probability of a false negative test for individuals exposed $a$ days ago, where the test is assumed to occur two days before departure, when the infection prevalence at $i$ is $n_i(t,a)$.

It is assumed that individuals with symptoms who receive a false negative test result on a pre-departure test still travel. When no pre-departure test is required, some travelers do not travel due to symptoms. The number of infected travelers that were exposed $a$ days ago and arrive from origin $i$ at $t$ when no pre-departure test is required is,

$$\mathrm{T}_i^{k,0}(t,a) = v_i^k(t) n_i(t,a) \rho \psi \Lambda(a), \tag{D9}$$

where travelers develop symptoms with probability $\rho$, travel irrespective of symptoms with probability $\psi$, and first develop symptoms before departure (when their infection age is $a$ at departure) with probability $\Lambda(a)$, where

$$\Lambda(a) = \sum_{\alpha=0}^{a} \lambda(\alpha) \tag{D10}$$

and,

$$\lambda(a) = \Gamma(a, \text{shape} = 5.807, \text{scale} = 0.948)\Delta a. \tag{D11}$$

This parameterization is from Lauer et al (2020) and corresponds to the first symptoms occurring a mean of 5.5 days after exposure with a standard deviation of 2.3 days (Hart et al, 2021). The probability density is discretized with $\Delta a = 1$ day because other data for our model occurs only at 1 day intervals. The cumulative mass function (equation D10) is used because travelers with infection age $a$ that decided not to depart on date $t$ may have first developed symptoms at any time 0 to $a$ days after exposure.

In Canada, a pre-arrival test policy for international travelers was enacted on January 7, 2021. Having a negative COVID-19 test result (72 hours prior to departure) was required for travelers departing from international ports (Government of Canada, 2020) and from May 15, 2021 this was necessary for all travelers (Traveler COVID-19 Testing 2021, see Table D6). The number of infected travelers arrived in NL on day $t$ with an infection of age $a$ is,

$$T_i^k(t,a) = \mathbb{1}_{\text{PRE,i}}^k(t)\ T_i^{k,1}(t,a) + (1 - \mathbb{1}_{\text{PRE,i}}^k(t))\ T_i^{k,0}(t,a), \tag{D12}$$

where $\mathbb{1}_{\text{PRE,i}}^k(t)$ is an indicator variable that is equal to 1 if a pre-departure test is required for travelers of type $k$, departing from origin $i$ at time $t$, and 0 otherwise.

#### D.3 Mean time to developing symptoms after arrival

The mean time to developing symptoms after arrival is calculated as,

$$\bar{a} = \sum_{a=0}^{13} \sum_{a_t=0}^{13} a\lambda(a + a_t) p(a_t), \tag{D13}$$

where the probability that an arriving traveler has age of infection $a_t$ at arrival is,

$$p(a_t) = \frac{\frac{1}{14}\left(1 - \rho(1-\psi)\Lambda(a)\right)}{\sum_{a=0}^{13} \frac{1}{14}\left(1 - \rho(1-\psi)\Lambda(a)\right)}. \tag{D14}$$

Since we only predict cases each day, and $\gamma(a)$ is a discrete function defined each day, $\bar{a}$ from equation D13 is rounded to the nearest integer before being used in equation D16.

The probability of infections of age $a_t$ at arrival, amongst arriving travelers is calculated assuming that exposure times for travelers are uniformly distributed as 0 to 13 days before departure, and that $1-\psi$ travelers that develop symptoms before departure do not travel. The average timing of first symptoms after arrival is calculated assuming no pre-departure test. As some travelers do not travel due to symptoms, the distribution of ages of infection in arriving travelers is skewed right (i.e., the tail is to the right) because more arriving travelers were exposed shortly before departure.

The use of $\bar{a}$ in equation D16 is an approximation because, more exactly, a traveler arriving on day $t$ with infection age $a$ might first show symptoms on any of the days post-arrival (14 possible days). For our model, there are 273 possible arrival days (September 1, 2020 to May 31, 2021) and 14 possible ages of infection at arrival, which means that it is necessary to calculate 3,822 values corresponding to each arrival date and age of infection. If we consider that travelers might first develop symptoms on any of the 14 days they are in self-isolation after arrival, then the number of calculations necessary increases to 53,508. To reduce the number of calculations needed, we assume that all arriving travelers first experience symptoms $\bar{a}$ days after arrival, where the value of $\bar{a}$ has some epidemiological basis (see equation D13), but is assumed equal for all travelers irrespective of when they were exposed.

#### D.4 Post-arrival testing

Federal and provincial testing requirements during the study period are summarized in Table D6. Passengers on flights that were subject to exposure notifications were also required to complete PCR tests. These notifications are listed in Table D7.

**Table D6** COVID-19 testing requirements for travellers arriving in Newfoundland and Labrador due to border measures implemented by the Government of Canada or Special Measures Orders issued by Newfoundland and Labrador.

| Dates | Test policy |
|---|---|
| 2020-09-09 | Asymptomatic rotational workers are exempt from the requirement to self-isolate for 14 days if they receive a negative test result for a test completed between five and seven days after their return |
| 2020-11-25 | Rotational workers required to wait until day seven of their 14-day self-isolation period to arrange COVID-19 testing |
| 2021-01-07 | Requirement for mandatory negative COVID-19 test result for air passengers entering Canada (Federal measure) |
| 2021-03-12 | Domestic rotational workers can cease self-isolation if testing negative (tested upon return, day 7 and on day 11,12 or 13) |
| 2021-03-27 | Updated rotational worker requirements: must isolate away from families for 14 days, cannot avail test on day 7 or modified self-isolation |
| 2021-04-19 | Essential workers entering the province required to self-isolate until they receive their 1st negative test result |
| 2021-05-15 | COVID-19 testing order for individuals arriving in NL (testing requirements during their 14-day self-isolation) |

PCR test sensitivity is based on Figure 3B in Hellewell et al (2021). As such,

$$\gamma(a) = [0,\ 0.05,\ 0.1,\ 0.55,\ 0.78,\ 0.77,\ 0.73,\ 0.68,\ 0.64,\ 0.59,\ 0.55,\ 0.49,\ 0.43,\ 0.37,\ 0.31], \tag{D15}$$

for infection ages $a = [0, 1, \ldots, 14]$ days.

The mechanistic model requires information on days since exposure for infected travelers as this affects the PCR test sensitivity, and the timing of symptoms post-arrival, which affects when test results are reported. We assume that travelers that test positive due to an exposure notification were infected on their flight. For other travelers, we assume that the exposure time is uniformly distributed between 0 and 13 days prior to departure. This range was selected because any traveler infected more than 13 days prior to departure is unlikely to be infectious after arrival (Ferretti et al., 2020). We assume that a fraction of infected travelers originating from Canada and internationally, and all travelers that test positive on pre-departure tests, do not travel, which impacts the distribution of days since exposure for infected travelers, given that they have arrived in NL.

Regular travelers: symptomatic after arrival. Regular travelers (i.e., travelers that are not rotational workers) that departed from $i$, may have been reported as a travel-related case in NL on day $t$, if they developed symptoms post-arrival and requested a PCR test. The predicted number of such travel-related cases is,

$$R_i^{r,s}(t) = \rho\sigma \sum_{a=0}^{13} T_i^r(t - \bar{a} - t_r - t_{rep}, a)\ \gamma(a + \bar{a} + t_r), \tag{D16}$$

**Table D7** Exposure notification from Public Health Advisories in NL during the COVID-19 pandemic where passengers on arriving flights were asked to arrange COVID-19 testing (Government of Newfoundland Labrador, 2023).

| Date | Exposure notification to passengers | Origin | Destination |
|---|---|---|---|
| 2020-09-27 | WestJet Flights 306 arrived on Monday, September 21<br>WestJet Flights 328 arrived on Monday, September 21 | Winnipeg, MB<br>Toronto, ON | St. John's, NL<br>St. John's, NL |
| 2020-10-05 | Air Canada Flight AC604 arrived on Wednesday, September 30<br>Air Canada Flight AC8876 arrived on Wednesday, September 30 | Toronto, ON<br>Halifax, NS | Halifax, NS<br>Deer Lake, NL |
| 2020-10-23 | Air Canada Flight 7484 arrived on Monday, October 12 | Toronto, ON | Deer Lake, NL |
| 2020-11-04 | Air Canada Flight 7484 arrived on Friday, October 30 | Toronto, ON | Deer Lake, NL |
| 2020-11-23 | Air Canada Flight 8880 arrived on Thursday, November 19 | Halifax, NS | Deer Lake, NL |
| 2020-11-24 | People who returned to NL from Nova Scotia in the last two weeks, and who visited bars in Halifax and surrounding metro communities | NS | NL |
| 2020-12-04 | WestJet Flight 3428 arrived on Thursday, November 26 | Halifax, NS | St. John's, NL |
| 2020-12-20 | Several flight advisories this weekend:<br>Air Canada Flight 8862 arrived on Monday, December 7<br>Air Canada Flight 690 arrived on Friday, December 11<br>Air Canada Flight 8862 arrived on Friday, December 11<br>Air Canada Flight 8862 arrived on Tuesday, December 15<br>Air Canada Flight 690 arrived on Tuesday, December 15<br>Air Canada Flight 690 arrived on Thursday, December 17 | <br>Halifax, NS<br>Toronto, ON<br>Halifax, NS<br>Halifax, NS<br>Toronto, ON<br>Toronto, ON | <br>Gander, NL<br>St. John's, NL<br>Gander, NL<br>Gander, NL<br>St. John's, NL<br>St. John's, NL |
| 2020-12-29 | Air Canada Flight 8880 arrived on Tuesday, December 22 | Halifax, NS | Deer Lake, NL |
| 2021-01-20 | Who traveled on the MV Blue Puttees to and from North Sydney, Nova Scotia, and Port Aux Basques between Tuesday, December 29 and Saturday, January 16 | Sydney, NS | Port Aux Basques, NL |
| 2021-02-15 | Air Canada Flight 7484 arrived on Thursday, February 11, | Toronto, ON | Deer Lake, NL |
| 2021-03-04 | Air Canada Flight 8996 arrived on Thursday, February 25 | Halifax, NS | St. John's, NL |
| 2021-04-19 | Air Canada Flight 8008 arrived on Monday, April 13 | Toronto, ON | Deer Lake, NL |
| 2021-05-02 | Air Canada Flight 8016 arrived on Friday, April 30 | Montreal, QC | St. John's, NL |
| 2021-05-06 | Air Canada Flight 7540 arrived on Tuesday, May 4 | Toronto, ON | Deer Lake, NL |
| 2021-05-11 | Air Canada Flight 7542 arrived on Monday, May 10 | Toronto, ON | Deer Lake, NL |

where $T_i^r(t - \bar{a} - t_r - t_{rep}, a))$ is the number of infected regular travelers arriving from $i$, with an infection of age, $a$, where $\bar{a} + t_r + t_{rep}$ is the number of days after the travellers arrival when the travel-related case would be reported, $\rho$ is the probability that infected travelers have symptomatic infections, $\sigma$ is the probability that travelers with symptoms request a test, and $\gamma(a)$ is the probability of a positive test result for an infection that is $a$ days since exposure. The mean time to develop symptoms after a traveler's arrival is $\bar{a}$ days (see section D.3 in the Supplementary information for the derivation). The time between requesting a PCR test and the test being performed is $t_r$ days, and the time between the test and reporting of the results is $t_{rep}$ days.

Regular travelers: exposure notification. Regular travelers that departed from $i$, may have been reported as a travel-related case in NL on day $t$, if they were asked to complete PCR testing due to an exposure notification on their arriving flight. For the arrival dates corresponding to exposure notifications (see Table D7 in the Supplementary Information), the predicted number of such travel-related cases is,

$$R_i^{r,e}(t) = v_i^r(t - t_e - t_r - t_{rep})\phi_1\ \phi_2\ \gamma(t_e + t_r), \tag{D17}$$

where $v_i^r(t - t_e - t_r - t_{rep})$ is the number of regular travelers arriving from $i$, the number of days after the travelers' arrival when the travel-related case would be reported is $t_e + t_r + t_{rep}$, the number of days after arrival when the exposure notification is issued is $t_e$, $\phi_1$ is the probability of being infected on a flight, and $\phi_2$ is the proportion of flights arriving on day $t$ that were included in the exposure notification.

Equation (D17) assumes that travelers who were exposed on flights were not infected prior to departure. While this many not always be the case, with few exposure notifications it is unlikely that our equations will result in substantial double counting of infected individuals because we counted these individuals both as infected during a flight, and infected pre-departure.

Rotational workers: mandatory testing. From the early stages of the pandemic, in NL there were specific post-arrival testing measures that applied to rotational workers (see Supplementary Information, Table D6). Rotational workers were required to complete up to 3 post-arrival PCR tests. We define the number of rotational workers that test positive on their first post-arrival test occurring $t_1$ days after arrival from origin $i$, as,

$$R_i^{rw,1}(t) = \sum_{a=0}^{13} \gamma(a+t_1)\; T_i^{rw}(t - t_1 - t_{rep}, a), \tag{D18}$$

where $T_i^{rw}(t - t_1 - t_{rep}, a)$ is the number of infected rotational workers arriving from $i$, with an infection of age, $a$, where $t_1 + t_{rep}$ is the number of days after the travelers' arrival when the travel-related case would be reported.

For positive results on the second and third post-arrival tests, occurring $t_2$ and $t_3$ days after arrival, but not any post-arrival tests prior (i.e., for the third post-arrival test, but not testing positive on either the first or second post-arrival test) is,

$$R_i^{rw,2}(t) = \sum_{a=0}^{13} \gamma(a+t_2)\; T_i^{rw}(t - t_2 - t_{rep}, a)\; (1-\gamma(a+t_1))\; T_i^{rw}(t - t_2 - t_{rep}, a) \tag{D19}$$

$$R_i^{rw,3}(t) = \sum_{a=0}^{13} \gamma(a+t_3)\; T_i^{rw}(t - t_3 - t_{rep}, a)\; (1-\gamma(a+t_1))\; (1-\gamma(a+t_2))\; T_i^{rw}(t - t_3 - t_{rep}, a). \tag{D20}$$

The predicted number of travel-related cases reported due to positive test results from rotational workers is calculated by summing the number of positive results for first, second, and third post-arrival tests (where applicable) that would be reported on day $t$,

$$R_i^{rw}(t) = \sum_k R_i^{rw,k}(t). \tag{D21}$$

We do not consider tests for rotational workers based on developing symptoms when at least one post-arrival test was mandatory for rotational workers.

### Appendix E Additional details regarding the results

This section provides additional details concerning the results reported in the main text.

#### E.1 Estimated parameters for the best models

The best model for travel-related cases of Canadian origin, $Y_t$ is,

$$\eta_t = \beta_0 + \sum_i \beta_i X_{i,t-5} \tag{E22}$$

$$y_t = \exp(\eta_t) \tag{E23}$$

$$Y_t \sim \text{POISSON}(y_t) \tag{E24}$$

where $y_t$ is the number of travel-related cases predicted by the statistical model on day $t$, and $X_{i,t-5}$ is infection prevalence, travel volume (indicated with $v$ in the variable names below), or the product of travel volume and infection prevalence for the origin $i$ on day $t-5$. A delay of five days was selected because our mechanistic model parameterization was that on average travelers would develop symptoms one day after arrival, would be tested three days after developing symptoms, and would be reported one day later. The estimates of the coefficients are $\beta_0 = -0.4$ (-3.6, 2.6) and $\beta_i$ for each $i$ are: AB: 5407* (2120,8695), vAB: 0.02 (-0.002, 0.04); BC: -1605* (-31732, -957); vBC: -0.009 (-0.06, 0.04); SK: -3270 (-8374,1121); vSK: -0.02 (-0.3, 2.0); MB: 1702* (458,2939), vMB: 0.05 (-0.1,0.2); ON: 4115 (-14671, 22544); vON: -0.02 (-0.04, 0.01); QC: -5335 (-16151,5105); vQC: -0.04 (-0.1,0.02); NB: -38080 (-93374, 18287); vNB: -0.04 (-0.09, 0.02); NS: 5509 (-10950,22462), vNS: 0.001 (-0.006,0.008); PEI: 11730 (-33071,55); vPEI: 0.07 (-0.01, 0.1), TR: 3468 (-4615, 10837); vTR: 0.008 (-0.015,0.03); AB×vAB: -47* (-82, -13); BC×vBC: 306 (-106,718); SK×vSK: 498 (-549,1611); MB×vMB: 2 (-183,182); ON×vON: 11.4 (-128,152);

QC×vQC: 125 (-286,541); NB×vNB: 230400 (-958,5203); NS×vNS: 10 (-192,205); PEI×vPEI: 657 (-7129,16981); and TR×vTR: -56 (-348,224) where the asterisk indicates estimates that are significantly different than zero and 95% confidence intervals are provided in the parentheses.

The best model for travel-related cases of international origin, $Y_t$ is,

$$\eta_t = \beta_0 + \beta X_{i,t-5} \quad \text{(E25)}$$
$$y_t = \exp(\eta_t) \quad \text{(E26)}$$
$$Y_t \sim NEGBIN(Y_t, \theta) \quad \text{(E27)}$$

where $X_{i,t-5}$ is infection prevalence, travel volume (indicated with $v$ in the variable name), or infection prevalence times travel volume in each of eight countries, $i$, with a significant amount of travel to NL lagged by five days to reflect reporting delays. Parameter estimates are $\beta_0 = -3.2^*$ (-5,-1.3) and the $\beta_i$ values for each country are: France: -135 (-1153, 745); vFrance: -0.01 (-0.07,0.03); Guyana: -55 (-753,571), vGuyana: 0.08 (-0.02, 0.2); India: 427* (196,422); vIndia: -0.05 (-0.08,0.04); Norway: -128 (-5402,4614); v.Norway: -0.2 (-0.4,0.003); Philippines: -679 (-1958,409); vPhilippines: 0.04 (-0.08,0.2); Spain: -34 (-549,484); vSpain: 0.009 (-0.3,0.1); UK: -630 (-1912,698); vUK: -0.2 (-0.1,0.008); USA: 1965 (-726,4665); vUSA: 0.001 (-0.02,0.02); vOther Int: 0.01* (-0.004,0.03); France×vFrance: 65 (-22,165); Guyana×vGuyana: -73 (-170,13); India×vIndia: -7 (-27,11); Norway×vNorway: 837 (-135,1896); Philippines×vPhilippines: -0.5 (-147,41); Spain×vSpain: -281 (-1055,161); UK×vUK: 35 (-25,103); US×vUS: -0.9 (-20,19); and OtherInt×vOtherInt: -15 (-41,9.6) where * indicates values that are significantly different from zero and 95% confidence intervals are provided in the parentheses. The dispersion parameter for the negative binomial is estimated to be $\theta = 0.742$ with a standard error of 0.325.

We also considered models with a Poisson error distribution (Table E8), but because we assessed the best models to have the smallest negative log likelihoods, and the Poisson regressions fitted fewer parameters, the models with the negative binomial error distributions presented in the main text were assessed as better models. The only exception is the model described by equation which could not be fit using the negative binomial error distribution. The `glm.nb` function did not converge because the overdispersion parameter could not be estimated, but the other estimated model parameters were the same as reported for the Poisson error distribution under equation .

**Table E8** Identical to Table 3 but these models assume a Poisson error distribution

| Canada | nLL | LR | K | AICc |
|---|---|---|---|---|
| Travel volume × infection prevalence (provinces) | 244.3 | 325.3 | 31 | 559.1 |
| Infection prevalence (provinces) | 273.4 | 267.1 | 11 | 569.9 |
| Travel volume (provinces) | 318.6 | 176.8 | 11 | 660.2 |
| Travel volume × infection prevalence (aggregated) | 358.4 | 97.1 | 4 | 725.1 |
| Travel volume (aggregated) | 379.4 | 55.1 | 2 | 762.9 |
| Infection prevalence (aggregated) | 391.3 | 31.4 | 2 | 786.6 |
| Constant only | 407.0 | 0 | 1 | 816.0 |
| Without travel-related cases | 538.2 | not nested | 0 | 1076.5 |
| International | | | | |
| Travel volume × infection prevalence (countries) | 140.3 | 76.9 | 27 | 340.8 |
| Infection prevalence (countries) | 159.1 | 39.3 | 9 | 336.8 |
| Travel volume × infection prevalence (aggregated) | 165.1 | 27.3 | 4 | 338.3 |
| Infection prevalence (aggregated) | 165.5 | 26.3 | 2 | 335.1 |
| Travel volume (countries) | 171.2 | 14.9 | 10 | 363.4 |
| Travel volume (aggregated) | 177.5 | 2.4 | 2 | 359.4 |
| Constant only | 178.7 | 0 | 1 | 359.4 |
| Without travel-related cases | 191.2 | not nested | 0 | 382.4 |

#### E.2 More models representing data gaps

We formulated statistical models that represent other types of data gaps to understand how these gaps might affect the reliability of importation models.

##### E.2.1 Only one source of travel volume data

We formulated models that consider only one source of travel volume data. The total travel volume during the time that travel-related cases are reported depends substantially on the TDF data, so we consider models formulated using the IATA and FC data sources only, which are compared to models formulated with total travel volume (which is the quantity that is used throughout our other analyses). Table 1 describes the limitations of the IATA and the FC data. Notably, both these sources only report monthly travel volumes, so models developed for these data sources need to be fit to data describing monthly travel-related cases. Further, the IATA data reports travel volumes before the pandemic, and the FC data only reports international arrivals. For the IATA data, we used the travel volumes for 2019 to predict travel-related cases for the same month during the pandemic: September 2020 - May 2021. For the FC data, we only predicted travel-related cases of international origin.

Results of the modelling that considers single sources of travel volume data are in Table E9. These models are not nested so we assess them based on the AICc sources. These AICc scores do not suggest that any of the models are better than the others, including the constant only model. Therefore, our ability to test models using this approach is limited by needing to aggregate the travel-related cases by month, which reduces the number of observations to just nine (as compared to 273 observations of daily travel-related cases each of Canadian or international origin). The interaction models contained only the interaction term

**Table E9** Models for monthly travel-related cases reported in NL from Canadian and international origins considering different types of travel volume data. All Canadian provinces are aggregated as there are only nine observations, so the data are not sufficient to support the estimation of ten coefficients.

| Canada | nLL | K | AICc | Δ AICc |
|---|---|---|---|---|
| Constant only | 38.2 | 2 | 82.4 | 0.0 |
| Total travel volume | 36.3 | 3 | 83.4 | 1.0 |
| IATA only | 37.6 | 3 | 85.9 | 3.5 |
| Total travel volume × infection prevalence | 37.7 | 3 | 86.3 | 3.9 |
| IATA × infection prevalence | 37.9 | 3 | 86.5 | 4.1 |
| International | | | | |
| Constant only | 25.5 | 2 | 57.1 | 0.0 |
| IATA × infection prevalence | 24.7 | 3 | 60.2 | 3.1 |
| Frontier counts only | 24.8 | 3 | 60.4 | 3.3 |
| IATA only | 25.0 | 3 | 60.7 | 3.6 |
| Total travel volume | 25.0 | 3 | 60.9 | 3.9 |
| Total travel volume × infection prevalence | 25.4 | 3 | 61.7 | 4.3 |
| FC × infection prevalence | 25.5 | 3 | 61.8 | 4.4 |

##### E.2.2 Infection prevalence not corrected for underreporting

We also considered models with infection prevalence that was not corrected for underreporting. Table E10 shows that correcting for under-reporting may improve models, particularly when travel-related cases of Canadian origin.

**Table E10** Models for daily travel-related cases reported in NL from Canadian and International origins considering infection prevalence corrected for underreporting (corrected? = yes) and not corrected for underreporting (corrected? = no). Explanatory variables are stratified for origin from each non-NL Canadian province or the territories and eleven countries with non-negligible travel to NL.

| Canada | corrected? | nLL | K | AICc | Δ AICc |
|---|---|---|---|---|---|
| Total travel volume × infection prevalence | no | 242.4 | 32 | 557.7 | 0.0 |
| Total travel volume × infection prevalence | yes | 244.3 | 32 | 561.7 | 4.0 |
| Infection prevalence | yes | 270.8 | 12 | 566.8 | 9.1 |
| Infection prevalence | no | 283.4 | 12 | 592.0 | 34.3 |
| Constant only | | 333.2 | 2 | 670.5 | 112.8 |
| International | | | | | |
| Infection prevalence | no | 142.1 | 10 | 305.1 | 0.0 |
| Infection prevalence | yes | 143.0 | 10 | 306.8 | 1.7 |
| Constant only | | 152.7 | 2 | 309.4 | 4.3 |
| Total travel volume × infection prevalence | yes | 131.4 | 28 | 325.7 | 20.6 |
| Total travel volume × infection prevalence | no | 137.0 | 28 | 336.8 | 31.7 |

### Appendix F An alternative version of the mechanistic model

The mechanistic model does not fit the data well (Figure 2E,F) because it is not fit to the travel-related case data. The mechanistic model can fit the data much better if we set the under-reporting coefficients to be ten times larger than their values in Table C4. The improved model fit is shown in Figure F4.

Additionally, we note that ‘spikes’ can occur in the mechanistic model predictions due to exposure notifications that require travelers to get tested. Such ‘spikes’ do not occur for the statistical models as these do not consider exposure notifications.

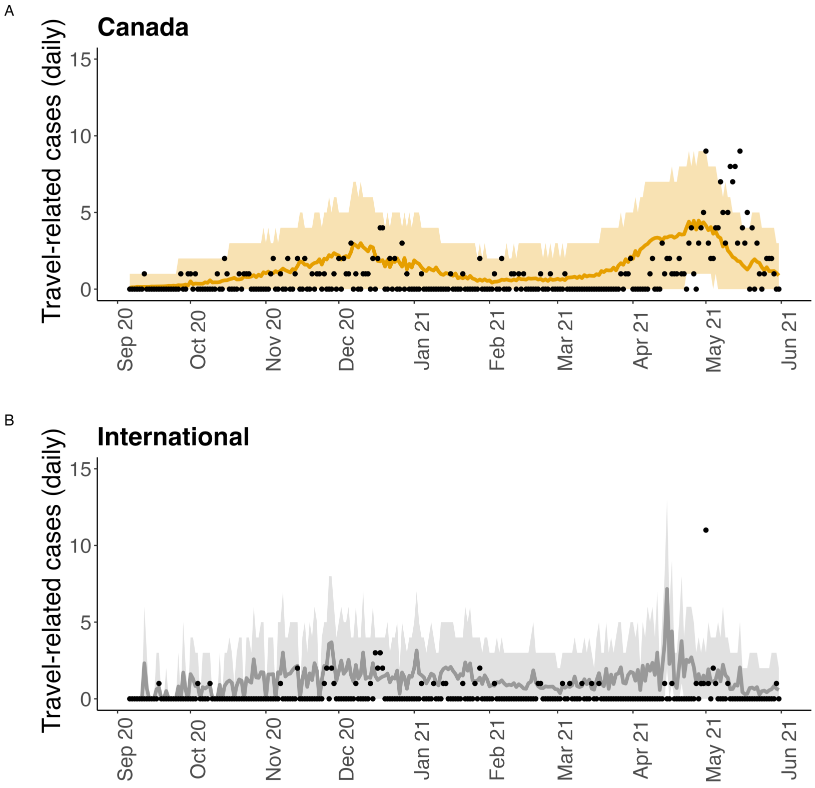


**Fig. F4** The mechanistic model (i.e. as shown in Figure 2E,F) when the underreporting coefficients are multiplied by 10.
